# Analytical and Clinical Validation of a High Accuracy Fully Automated Digital Immunoassay for Plasma Phospho-Tau 217 for Clinical Use in Detecting Amyloid Pathology

**DOI:** 10.1101/2024.10.31.24316186

**Authors:** David Wilson, Meenakshi Khare, Gallen Triana-Baltzer, Michele Wolfe, Patrick Sheehy, Karen Copeland, Lyndal Hesterberg, Ann-Jeanette Vasko, Wiesje M. van der Flier, Inge M.W. Verberk, Charlotte E. Teunissen, Mike Miller

**Affiliations:** Quanterix Corporation, Billerica, MA, USA; Johnson and Johnson Innovative Medicine, La Jolla CA, USA; Boulder Statistics, Steamboat Springs, CO; HCS, Inc, Denver, CO; Neurochemistry Laboratory, Department of Laboratory Medicine, Amsterdam UMC, Vrije Universiteit Amsterdam, Amsterdam Neuroscience, Amsterdam, The Netherlands

**Author notes:** Address correspondence to: David H Wilson, PhD.

**Keywords:** Digital immunoassay, biomarker, p-Tau 217, amyloid, Alzheimer’s disease, validation

## Abstract

**Objectives:** Phospho-tau 217 (p-Tau 217) in plasma has emerged as a promising biomarker for detecting amyloid pathology in plasma as an aid in diagnosis of Alzheimer’s disease. Because of its low abundance in plasma, reliable quantitation in blood is challenging. Digital single molecule array (Simoa) technology provides higher analytical sensitivity than possible with conventional analog immunoassays, enabling precise measurement of plasma p-Tau 217. We report the analytical and clinical validation of a Simoa digital immunoassay for p-Tau 217 across highly diverse clinical cohorts that meets consensus criteria for confirmatory test performance to aid the diagnosis of Alzheimer’s disease in people with cognitive impairment.

**Methods:** A Simoa p-Tau 217 assay utilizing a 2-cutoff approach was analytically validated using industry standard protocols, and diagnostic thresholds were clinically validated across 2 clinically diverse independent cohorts (total n = 873) using amyloid PET or CSF biomarkers as comparators.

**Results:** The assay exhibited acceptable analytical characteristics, including analytical sensitivity enabling measurement of plasma p-Tau 217 in all clinical samples and a lower limit of quantitation of 0.006 pg/ml. For samples not in the intermediate zone between the 2 cutoffs, the assay gave a clinical sensitivity of 90.3%, a specificity of 91.3%, and overall accuracy of 90.7%. 30.9% of the samples fell into the intermediate zone between the lower and upper cutoffs (0.04 and 0.09 pg/mL respectively). With an amyloid prevalence of 50%, typical for mild cognitive impairment, the positive and negative predictive values were 91.4% and 90.4% respectively.

**Conclusions:** The analytical characteristics are suitable for implementation of the Simoa p-Tau 217 assay as a lab developed test under the Clinical Laboratory Improvement Act (CLIA), and the clinical performance characteristics meet consensus criteria for a confirmatory plasma test to aid in Alzheimer’s diagnosis.

**RESEARCH IN CONTEXT:** *Evidence before this study:* Following rapid advances in plasma testing for AD-related biomarkers, the field has coalesced around p-Tau 217 as the most accurate single blood-based biomarker for detecting the presence of amyloid pathology. There have been a number of recent reports and conference presentations on different tests for plasma p-Tau 217 including large head-to-head comparisons. To date, however, none of these reports examine the analytical and clinical performance characteristics of the test--or the methodologies by which these characteristics were determined--in sufficient detail for a critical evaluation of the fitness of the test for its intended clinical use. Indeed, it has been the lack of robust analytical and clinical validation reporting that has led some to question if some of these tests are ready for prime time.

*Added value of this study:* Our work is the first to describe in detail the analytical and clinical performance characteristics of a high-accuracy laboratory developed test for plasma p-Tau 217 validated for clinical use. While the test is not currently FDA cleared, the validation studies were conducted with study protocols recommended by the FDA that generally exceed the standards required of LDTs developed and offered for clinical use under the Clinical Laboratory Improvement Act (CLIA). This report helps advance the field by describing the level of validation that is compatible with the FDA’s Final Rule (May 2024, FDA-2023-N-2177) in which LDTs were included as regulated in vitro diagnostic devices. Detailed validation reports like this can facilitate critical evaluation of whether a currently available LDT is fit for use as demonstrated by its validation robustness. More rigorous validation of these tests represents an important next step on the path toward widespread adoption and use of blood-based biomarkers for AD.

## INTRODUCTION

FDA approval of disease-modifying treatments (DMT) for Alzheimer’s disease (AD) and the likelihood of other potential approved DMTs in the pipeline highlights the urgent need for non-invasive widely available blood tests to facilitate timely diagnosis toward identifying patients eligible for treatment. Currently established biomarker-based approaches to diagnostic workup include positron emission tomography (PET) imaging and cerebrospinal fluid (CSF) biomarkers for amyloid and phosphorylated tau, both of which are invasive, expensive, and may not be widely available. Fortunately, significant advances have been made in recent years in the development of blood tests for detecting AD pathology driven by advances in high-sensitivity laboratory methods and high-quality antibody reagents. One such method, single molecule array (Simoa), enabled the introduction of high sensitivity immunoassays for numerous blood-based biomarkers of relevance in AD research and potentially diagnostics, including Abeta42/40, phosphorylated-tau isoforms, glial fibrillary acidic protein (GFAP) and neurofilament light (NfL) [1]. Among this slate of Simoa assays for plasma biomarkers, a high sensitivity assay for tau phosphorylated at the 217 residue (p-Tau 217) was described by Janssen R&D several years ago [2]. This assay was employed in numerous studies that have contributed to an important body of evidence highlighting this p-Tau isoform as generally outperforming two other well studied isoforms (p-Tau 181 and p-Tau 231) for detection of amyloid and tau pathology [3] and longitudinal monitoring and prognosis of disease progression [4]. Based on a sizable body of consistent evidence, a consensus has emerged that plasma p-Tau 217 represents the best single blood-based biomarker target currently available to aid in Alzheimer’s pathology detection. Reflecting this consensus, the Alzheimer’s Association (AA) Workgroup recently recommended plasma p-Tau 217 as the only blood-based biomarker that has demonstrated accuracy comparable to FDA-cleared CSF biomarker tests, enabling a confirmatory diagnostic use case with appropriate validation [5]. Such a use case has the potential to significantly attenuate reliance on PET and lumbar punctures for the AD diagnostic pathway. The proposed AA criteria also recommends that a blood test for plasma p-Tau 217 be designed with two cut-offs in recognition of signal overlap between diseased and non-diseased patients. The use of two cutoffs maximizes the negative and positive predictive values of the test and also yields a diagnostic ‘gray zone’ in which there is less certainty of amyloid status. The AA further recommended the plasma test should exhibit an accuracy of ≥90% for diagnostic use. Following closely behind the development of the AA guidance, the Us Against Alzheimer’s Global CEO initiative (CEOi) [6] similarly arrived at recommendations on confirmatory plasma test performance criteria and a 2-cutoff approach that mirrors that of the AA [7].

To address the need for a scalable high-accuracy blood test to facilitate AD diagnosis, we endeavored to validate a Simoa p-Tau 217 assay in accordance with CLIA standards and with sufficient clinical diversity and powering to establish robust diagnostic cutoffs for a lab developed test (LDT) for clinical use that meets AA and CEOi guidance for clinical performance. This paper describes the analytical and clinical validation of this Simoa plasma p-Tau 217 test, branded “LucentAD p-Tau 217” to indicate its commercial availability for clinical use.

## MATERIALS AND METHODS

### Apparatus

All Simoa p-Tau 217 assay testing was performed on the Simoa HD-X instrument, a fully automated digital immunoassay analyzer utilizing Simoa technology for isolation and counting of single molecules. The instrument pipettes sample directly from sample tubes or 96-well plates and processes immunoassays and data reduction with a steady state throughput of 66 tests/hour. Details of the instrument and its principles are given elsewhere [8].

### Assay principle and protocol

The single-molecule sensitivity of Simoa technology has been discussed [9]. In brief, Simoa is a digitized bead-based enzyme linked immunosorbent assay (ELISA) whereby diffusion of fluorescent reporter molecules at the signal step is constrained to 40-femtoliter wells in a microarray. By restricting diffusion to such a small volume, fluorophores generated by a single enzyme label can be detected in the array in 30 seconds. The arrays are composed of 216,000 wells which are counted simultaneously. Simultaneous counting of all femto wells enables rapid signal acquisition leading to rapid assays, which generally take 45-60 minutes for complete processing.

Through this simple digital processing approach, attomolar sensitivity can be obtained [9]. The Simoa p-Tau 217 assay design has been described [2]. In brief, the assay is a 3-step sandwich immunoassay in which sample is drawn from the sample tube by the instrument pipettor and mixed with anti-p-Tau 217 coated paramagnetic capture beads in a reaction cuvette. Following collection of the beads with a magnet, washing, and redispersion, biotinylated detector antibodies are combined with the beads and incubated. Following a second bead collection and wash, a conjugate of streptavidin-ß-galactosidase (SβG) is mixed with the capture beads for the third assay step. Following a third bead collection and wash, the capture beads are resuspended in a resorufin ß-D-galactopyranoside (RGP) substrate solution. Digital processing occurs when beads are transferred to the Simoa array disc [10]. Individual capture beads are sealed within microwells in the array. If p-Tau 217 has been captured and labeled, the ß-galactosidase hydrolyzes the RGP substrate into a fluorescent product that provides the signal for measurement. The concentration of p-Tau 217 in unknown samples is interpolated from a logistic 4-parameter standard curve. Time to assay completion per measurement is about an hour.

### Reagents

Four reagents were developed for the assay: paramagnetic p-Tau 217 capture beads, biotinylated detector, SβG conjugate, and sample diluent. The capture beads comprised a monoclonal anti-p-Tau 217 antibody (Janssen PT3, [11]) specific for an epitope spanning residues 210-220 with two phosphorylation sites (212 and 217) covalently attached by standard coupling chemistry to 2.7 μm carboxy paramagnetic microbeads (Agilent Technologies). The antibody-coated beads were diluted in Tris buffer with a surfactant and protein stabilizer (bovine). Biotinylated detector reagent comprised a monoclonal anti-tau antibody (Janssen HT43) specific for N terminal residues 7-20 that was biotinylated using standard methods and diluted in a PBS diluent containing surfactant and BSA. SβG was prepared by covalent conjugation of purified streptavidin (Thermo Scientific) and βG (Sigma) using standard coupling chemistry and in a phosphate buffer with a surfactant and protein stabilizer (bovine). Sample Diluent was formulated in PBS diluent with heterophilic blockers, EDTA, and a surfactant.

### Calibration

The assay is calibrated using purified peptide construct (MW 4929, New England Peptide, Gardner MA) composed of the N-terminal epitope (tau residues 7-20) and mid-region phosphorylated epitope (tau residues 210-220, phosphorylated at 212 and 217), connected by a 4-unit polyethylene glycol linker [12]. The peptides were HPLC purified, confirmed by mass spectral analysis, and the purified peptide mass-based concentration was determined by the manufacturer. Calibrators were prepared gravimetrically with nominal values of 0.002, 0.010, 0.039, 0.156, 0.625, 2.50, and 10.0 pg/mL based on volumetric dilutions and stored in phosphate buffer with a protein stabilizer (bovine), a surfactant, and ProClin 300 as a preservative.

### Analytical verification

Key assay analytical performance characteristics were verified at the Quanterix CLIA laboratory (Billerica, MA) in accordance with standard Clinical and Laboratory Standards Institute (CLSI) protocols. These studies verified performance across multiple instruments and reagent lots as indicated in Results.

#### Linearity

Assay linearity was tested according to CLSI Document EP06 Ed2 [13] with three replicates of 10 K2EDTA plasma samples distributed across the assay range to within 10% of the upper reportable limit. The 10 samples were prepared by admixing contrived elevated plasma samples with a native low p-Tau 217 plasma pool to arrive at evenly spaced p-Tau 217 concentrations across the range. Linearity was evaluated by linear regression analysis.

#### Sensitivity

Detection capability for limit of blank (LoB), limit of detection (LoD), and lower limit of quantitation (LLoQ) was estimated in accordance with CLSI Document EP17-A2 [14] across two reagent lots and a single HD-X instrument. For LoB, 20 replicates of the zero calibrator were assessed for each lot of reagents. LoB was estimated with the non-parametric analysis method across the two lots as prescribed in CLSI EP17-A2.

For LoD, 3 native plasma samples with low levels of p-Tau 217 and 3 contrived samples prepared by spiking antigen into the zero calibrator at low levels were tested in duplicate across 2 reagent lots. The pooled standard deviation (SD_L_) across the low-level samples was calculated according to EP17-A2 where LOD = LOB + Cp x SD_L_, where the multiplier Cp is given by

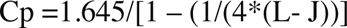

Here L = total number of all low-level sample results across all reagents and J = number of low-level samples.

For LLoQ, a set of 18 native plasma samples from healthy donors expected to have concentrations near the anticipated LLoQ were tested in duplicate over two runs, each with a different lot of reagents. For each lot, the precision profile (p-Tau 217 vs. replicate CV) was evaluated for the point at which the non-linear fit crossed the 20% CV level to define the LLoQ for that lot. The LLoQ was based on the worst performing lot of the 2 lots tested. Measurement accuracy was verified by confirming that the back calculation of the lowest p-Tau 217 calibrator (0.002 pg/mL, less than all the native samples) was within 80-120% of the expected concentration.

#### Repeatability and Reproducibility

Repeatability and within-laboratory precision were assessed according to CLSI document EP05-A3 [15] using 5 native plasma samples and a 5-day × 2 run × 2 replicate design across 2 reagent lots, 2 instruments, and 2 analysts (20 replicates/instrument, 40 total replicates). The 5 samples approximated low (near LLoQ), medium, and high levels of the assay measuring range. Two contrived plasma quality control samples (low and high, contrived with CSF from Alzheimer’s patients spiked into plasma) were also tested. Intra-assay repeatability was tested with 5 K2EDTA plasma samples from presumed normal donors, tested in replicates of 20 each. The average CV for each sample was then evaluated.

#### Specificity

Specificity of the assay was evaluated using synthetic tau peptides (Genscript Biotech, Piscataway, NJ) which included the N-terminal epitope and phosphorylation site epitopes at one of the following amino acid residues: 181, 205, 212/217 (positive control), 231, 231/235. Each peptide was prepared at 0.03, 0.3, 3.0, and 30 pg/mL in calibrator diluent and tested in replicates of three with one reagent lot and one instrument. Un-spiked buffer was used as a negative control.

#### Endogenous Interferences

Interference testing was performed according to CLSI EP07-Ed3 [16]. Three native K2EDTA plasma samples (one low, moderately positive, and one high positive, 0.024, 0.060, and 0.114 pg/mL respectively) were assessed for the impact of endogenous interferents (bilirubin, triglycerides, etc.) using one reagent lot and one instrument. Interferent stock solutions were prepared, where possible, at concentrations of at least 20 times the intended test concentration. Interferent stock solution was added to the test sample at a ratio of 1-part spiking solution stock to 19-parts sample. Equal volume of solvent used for the stock spiking solution (without interferent) was added to the control sample and care was taken not to dilute the matrix volume by more than 5%. In the case of total protein (human plasma albumin), the required amounts were directly weighed and added to the plasma samples. In case of human anti-mouse IgG (HAMA), a highly concentrated source was procured and diluted into the sample matrix to achieve the target concentration. All samples were tested in duplicate and all test and corresponding control conditions were performed in the same assay.

#### Sample Stability

The stability of 6 native K2EDTA plasma specimens (range: 0.024 – 0.114 pg/mL) was assessed at room temperature, refrigerated (2-8°C) and after 3 freeze-thaw cycles using one reagent lot and one instrument with guidance from CLSI document EP25-A Vol. 29 No. 20 [17]. Room temperature storage intervals were 4 and 8 hours, and the refrigerated storage intervals were 24 and 48 hours. -70°C storage served as the control condition.

#### Analytical Samples and Other Materials

To establish the detection capabilities of the p-tau 217 assay at the high end of the assay range, CSF from AD patients was used as a spiker into native K2EDTA samples. Similarly, endogenous quality control samples were prepared by spiking CSF from AD patients into commercially obtained K2EDTA plasma from individual presumed healthy donors.

### Clinical Validation

Clinical performance for classifying amyloid status was validated by comparison with either CSF biomarkers or amyloid PET across two independent cohorts: the Amsterdam Dementia Cohort (ADC) [18, 19] and the Bio-Hermes cohort [20]. Both cohorts were reviewed and approved by central or local ethics and safety review committees or boards. All participants (or their legally authorized representative) reviewed and signed an approved informed consent document to use medical data and biomaterials for research purposes.

#### Amsterdam Dementia Cohort

The ADC represents all patients who present to the Alzheimer Center Amsterdam of the Amsterdam University Medical Centers. These patients were referred for analysis of their cognitive complaints by their general practitioner or their local specialist. Each patient received the same standardized and multidisciplinary work-up which included history taking and cognitive examination by a neurologist, assessment of vital functions, informant-based history, and assessment of needs by a specialized dementia nurse, neuropsychological investigation, brain magnetic resonance imaging, electroencephalogram, standard laboratory work, and generally a lumbar puncture for CSF biomarker analysis. Some patients underwent amyloid PET scans instead of CSF collections. All patient cases were reviewed in a multidisciplinary meeting at which findings were reviewed toward arriving at a consensus on a diagnosis and treatment plan [18,19]. Diagnoses of Alzheimer’s dementia required an abnormal CSF biomarker profile or positive amyloid PET scan [21, 22]. Amyloid PET scans utilized either [^18^F]Florbetaben or [^18^F]Florbetapir and were classified as amyloid positive based on the presence of fibrillary amyloid pathology in the neocortex as evaluated by visual rating by a nuclear medicine physician. CSF Alzheimer’s biomarkers were measured with Roche Elecsys P-Tau 181/Abeta42 assays (510k K221842) using a cut point of 0.02 for amyloid positivity [23] or with Fujirebio Innotest ELISAs p-Tau 181/Aβ42 using a cut point of 0.06 [24]. Whole blood was obtained from each subject through vena puncture and processed into plasma by centrifuging at 1,800xg for 10 minutes at 20°C. Processed K2EDTA plasma samples were aliquoted in 0.5mL-portions in polypropylene tubes and stored at -80°C in the biobank until dry-ice transportation to the Quanterix CLIA lab for Simoa p-Tau 217 testing. The intended use population of the test is objectively impaired individuals. Accordingly, cases diagnosed with MCI (n = 123) and AD (n = 229) were chosen to comprise a portion (40%) of the training and validation cohorts. Details of these subgroups have been previously reported [24] and are shown in aggregate in Table S1. In addition, 50 each of cases diagnosed with frontal temporal dementia (FTD) and dementia with Lewy bodies (DLB) were examined. A proportion of these samples were also amyloid positive, and the accuracy of the test for detection of amyloid in these mixed pathology cases was characterized.

#### Bio-Hermes Cohort

From April 2021 through November 2022, 17 research sites prospectively recruited and enrolled consented study participants from their community-based populations. The clinical sites were recruiting centers for clinical trials investigating new drug treatments for Alzheimer’s. A key goal of the Bio-Hermes cohort was enrichment for ethnic/racial diversity. Participants who met inclusion criteria [20] were identified as belonging to one of the three clinical cohorts: cognitively unimpaired, MCI, and mild AD. Participants stratified to the MCI cohort met the following criteria: a diagnosis of MCI based on NIA-AA criteria [25] and verified through medical records, or had screening results as follows: MMSE score of 24 to 30 inclusive; RAVLT-delayed recall Score of at least 1 SD below the age-adjusted mean; and in the investigator’s judgment, minimal to mild functional impairment but with preservation of independence in functional abilities based on the FAQ score/study partner report. Participants stratified into the mild Alzheimer’s cohort met the following criteria: a diagnosis of probable Alzheimer’s based on the NIA-AA criteria [25] and verified through medical records, OR had screening results as follows: MMSE score of 20 to 24; RAVLT-delayed recall Score ≥1 SD below the age-adjusted mean; and in the investigator’s judgment, evidence of functional decline and dependence in functional abilities based on FAQ score/study partner report. Amyloid PET scans were obtained for all BioHermes participants following clinical diagnosis at designated imaging facilities near the recruitment sites. PET scans were conducted at a designated imaging facility near each site using [^18^F]Florbetapir tracer. For consistency of PET scan interpretations, all scans were uploaded into an imaging portal accessible for visual reading by IXICO Technologies Inc. Underrepresented population groups included Hispanics and non-Hispanic Blacks, overall constituted 27.8% of the symptomatic sub-cohort (MCI and mild Alzheimer’s). Whole blood samples were obtained from each subject through vena puncture in K2EDTA tubes, processed into plasma, and placed in the -80°C freezer within 4 hours. Samples were shipped on dry-ice to the Quanterix CLIA lab for Simoa p-Tau 217 testing. To meet the intended use population, cases diagnosed with MCI (n = 237) and mild AD (n = 284) were chosen to comprise a portion (60%) of the training and validation cohorts. Details of these subgroups have been previously reported [20] and are shown in aggregate in Table S1.

#### Diagnostic Threshold Development and Validation

To align with current recommendations for confirmatory plasma test performance [5, 7, 26], we endeavored to establish two cut points and achieve a minimum of 90% accuracy for the Simoa p-Tau 217 test on cohorts with objective cognitive symptoms. To do this, we utilized the samples from the MCI and AD groups within each cohort (ADC n = 352, Bio-Hermes n = 521) and randomized the samples of both cohorts combined into a training and validation sets stratified by MCI and AD status. The p-Tau 217 results from the validation were kept separate and blinded until use for validation. Diagnostic thresholds were modeled with the objective of achieving the accuracy target while minimizing the intermediate zone between the two cutoffs. The use of both cohorts for establishing the thresholds was deliberate to include the maximum diversity into the threshold setting. This diversity leads to the robustness of the thresholds in clinical practice.

#### Plasma Sample Analysis

Prior to analysis, K2EDTA plasma samples were thawed at room temperature for 60 minutes and centrifuged at 10,000 g for 10 minutes. Subsequently, p-Tau 217 concentrations from the clarified plasma supernatant were measured in duplicate on the Simoa HD-X analyzer in batches according to Quanterix CLIA laboratory SOPs using a single lot of reagents.

#### Statistical methods

Analytical study analyses followed the statistical techniques recommended in the appropriate CLSI guideline. Reporting of clinical performance metrics follow standard statistical practice, including effect sizes with 95% confidence intervals for key measures. P-values reported for comparisons of means are based on t-tests. P-values for comparisons of categorical variables are from likelihood ratio test of homogeneity. Clinical performance analysis was performed with JMP Pro 18.

To set clinical thresholds, a four factor, 40-run space filling design was used to model p-Tau 217 performance across false positive and false negative rates. The four design factors were the scale and shape parameters of log-normal distributions for amyloid negative and amyloid positive samples with the factor ranges determined by the 25th and 75th percentiles of 500 simulated distributions. A prediction profiler with a desirability function was used to evaluate and optimize the predicted performance (sensitivity, specificity % in indeterminant zone) in terms of false positive and false negative rates. The optimal rates were then converted to p-Tau 217 thresholds based on the fitted log-normal distributions.

## RESULTS

### Analytical Performance

#### Dose response and linearity

Figure 1 shows a representative calibration curve across a 3-log range. The low background typical for Simoa digital immunoassays is highlighted in Figure 1A. Linearity, evaluated across descending ratios from 1.0 high sample:low sample (ratio of 1.0 equals 100% AD (high) pool) to 0.875, 0.75, 0.625, 0.5, 0.375, 0.25, 0.125, 0.0625 and 0 (i.e., 100% CN pool), is depicted in Figure 1B. The average bias from expected values across the admixtures was 4%, with no improvement in fitting accuracy using a polynomial instead of linear fit. Linear regression statistics are depicted in Figure 1B.

**Figure 1.**
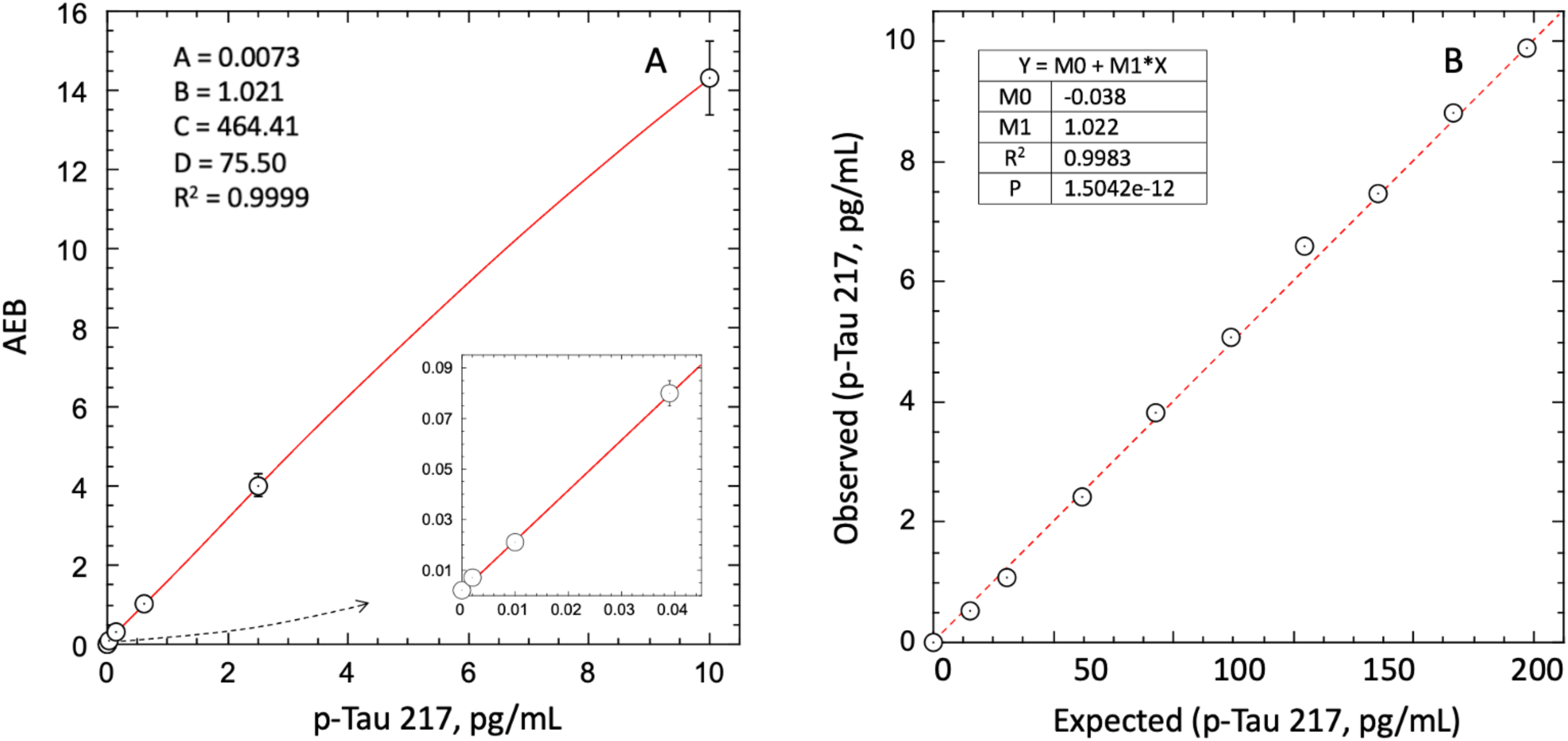
Dose response and linearity of Simoa LucentAD p-Tau 217 assay. (A), y-Axis reflects signal units (AEB). Fitting for optimal readback used 4PL regression (4parameters depicted). (B), Dashed line depicts linear fit.

#### Sensitivity

The highest LoB, LoD, and LLoQ results for the two reagent lots are reported for the assay. The highest LoB was determined to be 0.0005 pg/mL (0.5 fg/mL), and the highest LoD was calculated to be 0.0015 pg/mL. For LLoQ, precision profiles for repeated measurements of 18 native plasma samples from healthy donors are depicted in Figure 2. Most of the data exhibited less than 20% replicate CVs, hence with the reagent lot 1 data set and LLoQ could not be satisfactorily fit. Lot 2, however, gave a power fit that intersected the 20% CV threshold at 0.003 pg/mL. Correcting for a 1:2 pre-dilution of samples used in the instrument protocol, this yielded a functional LLoQ of 0.006 pg/mL. Accuracy of concentration readouts in this part of the assay range was verified by confirming readback of the lowest p-Tau 217 calibrator (0.002 pg/mL) was within 80-120%.

**Fig. 2.**
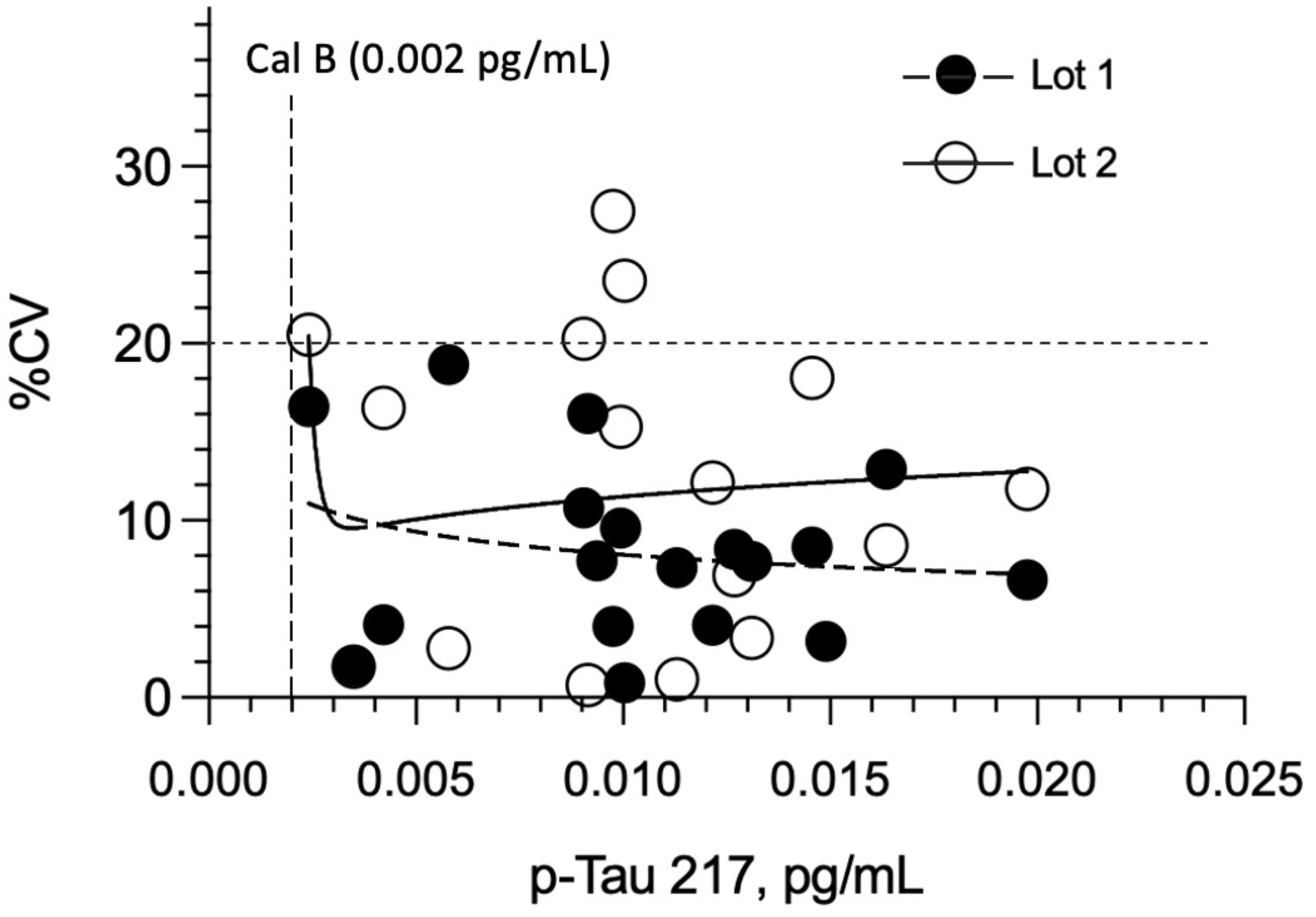
Concentration CVs of plasma from cognitively normal persons across reagent 2 lots. There was not a significant overall difference in imprecision between the lots (P=0.264). Analytical LLoQ was estimated where the power fit of Lot 2 data (solid line) met the 20% imprecision threshold (0.003 pg/mL). Concentrations depicted are uncorrected for 1:2 pre-dilution.

### Repeatability and Reproducibility

Repeatability and within-lab reproducibility for a panel of 6-8 amyloid negative and positive K2EDTA plasma samples spanning the lower and upper diagnostic cutoffs (as would be encountered in the intended use population) are summarized in Table 1. In both studies, percent coefficients of variation were ≤18%, even down to a level of 0.01 pg/mL, which is near the LLoQ and 4-fold lower than the lowest diagnostic cutoff (0.04 pg/mL, see Diagnostic Thresholds).

**Table 1:** Assay Repeatability and Reproducibility.

| **Intra Assay Repeatability (2 lots, 2 instruments, 2 analysts)** |  |  |  |
| --- | --- | --- | --- |
| **Sample** | **Mean (pg/mL)** | **SD (pg/mL)** | **%CV** |
| Plasma level 1 | 0.010 | 0.001 | 12.7 |
| Plasma level 2 | 0.101 | 0.011 | 11.2 |
| Plasma level 3 | 0.043 | 0.004 | 8.5 |
| Plasma level 4 | 0.052 | 0.003 | 5.7 |
| Plasma level 5 | 0.044 | 0.002 | 4.8 |
| Plasma level 6 | 0.060 | 0.005 | 8.0 |

| **Inter Assay Reproducibility (2 lots, 2 instruments, 2 analysts)** |  |  |  |
| --- | --- | --- | --- |
| **Sample** | **Mean (pg/mL)** | **SD (pg/mL)** | **%CV** |
| Plasma level 1 | 0.011 | 0.002 | 18.0 |
| Plasma level 2 | 0.113 | 0.012 | 10.6 |
| Plasma level 3 | 0.044 | 0.004 | 10.1 |
| Plasma level 4 | 0.050 | 0.004 | 8.6 |
| Plasma level 5 | 0.039 | 0.003 | 8.7 |
| Plasma level 6 | 0.059 | 0.009 | 15.8 |
| Plasma level 7 | 0.396 | 0.015 | 3.8 |
| Plasma level 8 | 0.136 | 0.021 | 15.5 |

#### Specificity

Figure 3 depicts the assay response to peptides phosphorylated at different amino acid residues. All p-tau peptides other than the peptide phosphorylated at the 212 and 217 residues peptide yielded a mean cross reactivity of <5%. The positive control (212/217) gave a recovery of 88.2% of the expected concentration.

**Figure 3.**
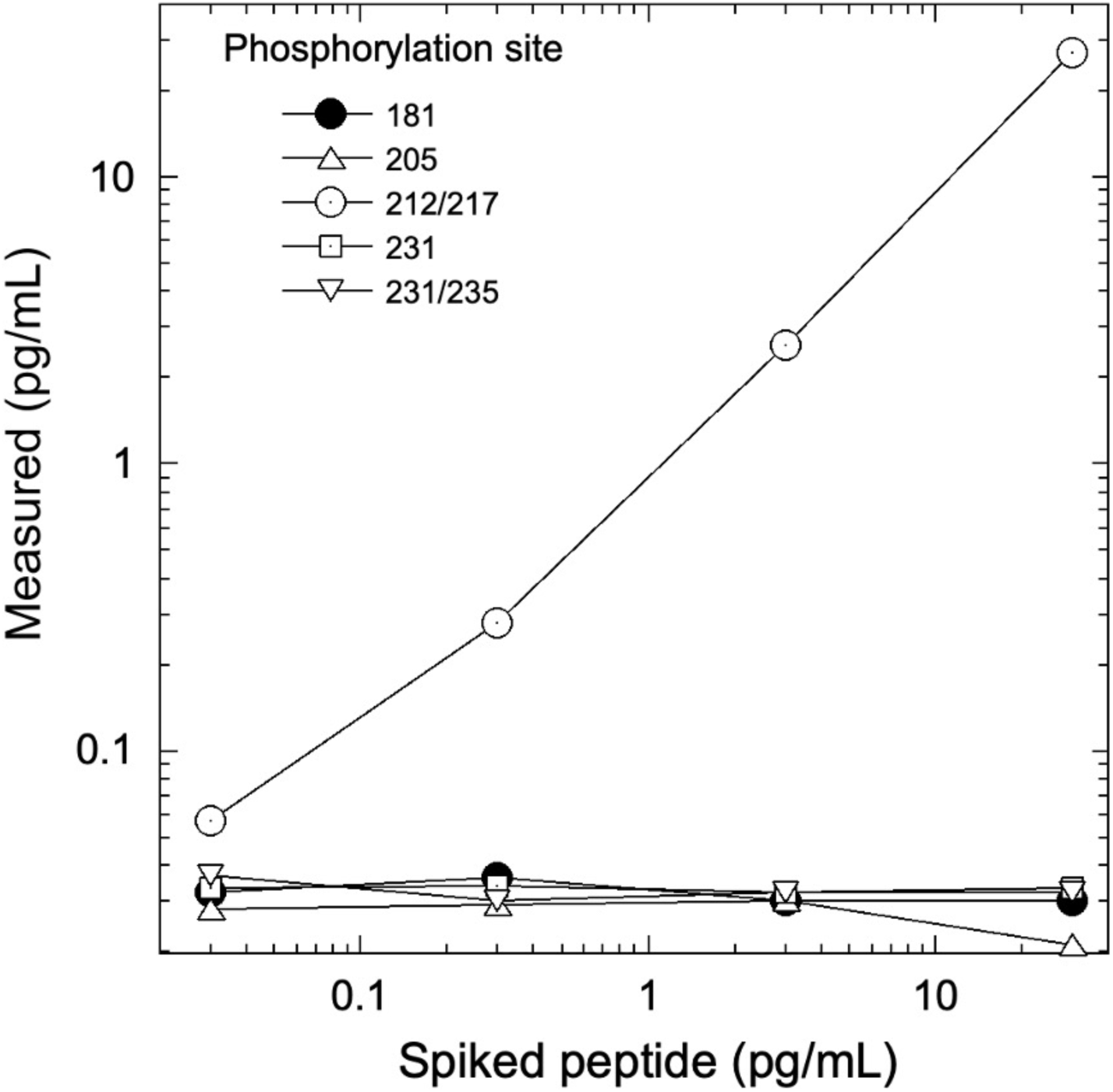
Assay response to tau peptides phosphorylated at different residues. Each peptide included an unphosphorylated N-terminal epitope along with 1 or 2 phosphorylated epitopes at the sites indicated. The assay was unreactive to sites other than 212/217. PT3 antibody reactivity to phosphorylation at the 212 vs. 217 sites was characterized previously [11].

#### Endogenous Interferences

Physiologically relevant levels of 8 potentially interfering endogenous substances (triglycerides, hemoglobin, total protein, conjugate and unconjugated bilirubin, HAMA, rheumatoid factor, and biotin) were tested by spiking into 3 plasma samples with p-tau 217 concentrations spanning the lower and upper diagnostic cutoff’s (0.04, 0.09 pg/mL respectively) as would be encountered in the intended use population. Table 2 exhibits the observed percent differences between spiked and un-spiked control samples, with overall mean % differences across the samples between -2.3% to 6.6%, with minimum and maximum % differences ranging from -11.8% to 15.8%.

**Table 2:** Endogenous Interferences.

| Potential Interferent | Level (Spiked) | Observed % difference between Spiked and Control specimen |  |  | Mean % Difference |
| --- | --- | --- | --- | --- | --- |
|  |  | Low Positive Plasma<br>(0.024 pg/ml) | Moderately Positive<br>Plasma (0.060 pg/mL) | High positive Plasma<br>(0.114 pg/mL) |  |
| Triglycerides | 1000 mg/dL | 5.4 | 4.3 | -7.4 | 0.8 |
| Hemoglobin | 500 mg/dL | 6.2 | 6.9 | 6.6 | 6.6 |
| Total Protein | 1g/dL | -11.8 | 0.0 | 4.8 | -2.3 |
| Conjugated Bilirubin | 20 mg/dL | 0.6 | 10.6 | 1.9 | 4.3 |
| Unconjugated Bilirubin | 20 mg/dL | 0.4 | 10.7 | 1.7 | 4.3 |
| HAMA | 10ng/mL | -2.0 | 15.8 | 6.1 | 6.6 |
| Rhumatoid Factor | 95 U/mL | 6.6 | 9.2 | -0.1 | 5.2 |
| Biotin | 0.360 mg/dL | -4.9 | 9.2 | -4.4 | 0.0 |

#### Sample Stability

p-Tau 217 as measured by the Simoa p-Tau 217 assay was found to be stable with a maximum average difference between test condition and control condition of 9% for 3 freeze-thaw cycles, 48 hours of refrigerated storage, and 8 hours of room temperature storage (Table 3).

**Table 3:** Stability of p-Tau 217 in EDTA plasma samples.

| Difference between test and control condition (pg/mL) |  |  |  |
| --- | --- | --- | --- |
| Condition | Average % | Lower 95% CI | Upper 95% CI |
|  | Difference |  |  |
| F/T 1 | 4.6 | -0.2 | 9.4 |
| F/T 2 | 2.1 | 0.2 | 4.0 |
| F/T 3 | 5.1 | -0.9 | 11.1 |
| 2-8°C, 24 hrs | 3.8 | 1.1 | 6.6 |
| 2-8°C, 48 hrs | 3.7 | -1.8 | 9.3 |
| RT, 4 hrs | 2.7 | -0.8 | 6.2 |
| RT, 8 hrs | 8.9 | 3.4 | 14.4 |

### Clinical Performance

#### Demographic and Clinical Characteristics

K2EDTA plasma samples from the ADC (n = 352) and study participants enrolled in the Bio-Hermes study (n = 521) were analyzed for p-Tau 217 and the results were compared with amyloid status by either CSF biomarkers or visual amyloid PET. The demographic and clinical characteristics of the two cohorts combined and separated by amyloid status are depicted in Table 4. Table S1 shows the demographics split by the original two cohorts. In the combined cohort (all data), the mean age was 70.1 (SD 8.0) years, with 50.3% female representation. However, the ADC reflected a younger population with a mean age of 65.4 years (SD 7.7, range 43-83), while the mean age in the Bio-Hermes cohort was 73.2 (SD 6.6, range 59-85) (Figure S1). Overall, the majority of the participants were white (86.6%), but 11.1% of study participants from Bio-Hermes were of black or African American origin. 13.1% of Bio-Hermes participants were Hispanic or Latino, with 27.8% of the participants in this cohort representing under-served minorities in total (including Asian, Pacific Islanders and Native Americans) (20). All individuals were symptomatic following the inclusion criteria of the study, with a diagnosis of either MCI (59%) or AD (including probable AD) (41%). 47.3% had one or more copies of apoE4 (APOE carriership). Overall, 56.7% of the participants were positive by either amyloid PET or CSF biomarkers. A breakdown of amyloid prevalence by subgroup is depicted in Figure 4. The prevalences differed significantly between the two cohorts. In the ADC, 56.3% of MCI subjects and by design >99% of the dementia patients were amyloid positive. The MCI prevalence reflects all comers to the Amsterdam tertiary care clinic, and the high prevalence among dementia subjects is due to selection of this clinical subgroup in which the diagnosis was confirmed by CSF biomarker results. On the other hand, 35.0% of the MCI subgroup in Bio-Hermes was amyloid positive, while 61.3% of the dementia subgroup was positive (Table S2). These comparatively lower numbers may reflect differing diagnostic criteria, use of recruited participants who had not been previously evaluated for cognitive symptoms, and clinical diagnoses being made prior to PET testing.

**Figure 4.**
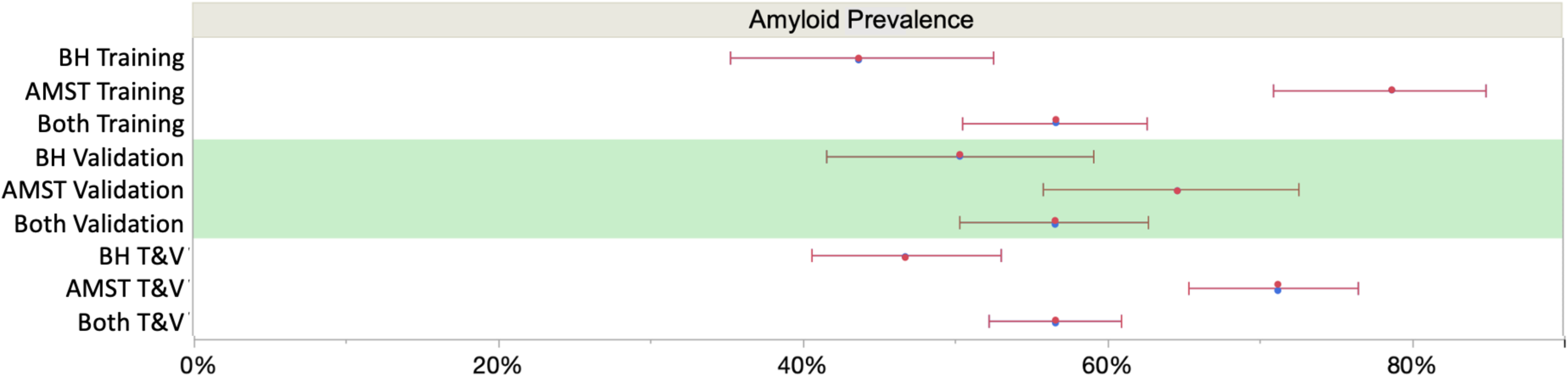
Subcohort prevalences (with 95% score proportion Cl’s). The overall mean prevalence was 57% across the two cohorts, which is skewed upward by the selected amyloid positives among the Amsterdam cohort. Assuming an intermediate prevalence of 50% [7], overall NPV and PPV were 90.4% and 91.4% respectively at the selected cutoffs (Figure 6). BH = Bìo-Hermes, AMST = Amsterdam Dementia Cohort.

**Table 4:** Demographic characteristics. Demographics of combined cohort. Amyloid classification based on amyloid PET or CSF biomarkers. Abbreviations: MCI = mild cognitive impairment; MMSE = Mini-Mental State Examination; PET = positron emission tomography.

|  | Aβ+ | Aβ- | All |
| --- | --- | --- | --- |
| **Age** |  |  |  |
| Mean (sd) | 70.4 (7.9) | 69.6 (8.2) | 70.1 (8.0) |
| Range | 45 - 85 | 43 - 85 | 43 - 85 |
| **Sex** |  |  |  |
| Female | 261 (52.7%) | 178 (47.1%) | 439 (50.3%) |
| **Race** |  |  |  |
| American Indian or Alaska Native | 0 (0.0%) | 1 (0.3%) | 1 (0.1%) |
| Asian | 5 (1.3%) | 5 (1.7%) | 10 (1.5%) |
| Black or African American | 18 (4.8%) | 42 (14.0%) | 60 (8.9%) |
| Native Hawaiian or other Pacific Islander | 0 (0.0%) | 1 (0.3%) | 1 (0.1%) |
| Other/Unknown | 10 (2.7%) | 9 (3.0%) | 19 (2.8%) |
| White | 343 (91.2%) | 243 (80.7%) | 586 (86.6%) |
| Missing†<br>‡Not included in %s | 119 | 77 | 196 |
| **Ethnicity** |  |  |  |
| Hispanic or Latino | 33 (13.52%) | 35 (12.64%) | 68 (13.05%) |
| Not Hispanic or Latino | 205 (84.02%) | 241 (87.00%) | 446 (85.60%) |
| Not Reported | 6 (2.46%) | 1 (0.36%) | 7 (1.34%) |
| Unknown (VUmc) | 251 | 101 | 352 |
| **Diagnosis** |  |  |  |
| AD | 122 (24.5%) | 1 (0.3%) | 123 (14.1%) |
| Probable AD | 147 (29.5%) | 93 (24.5%) | 235 (26.9%) |
| MCI | 229 (46.0%) | 286 (75.3%) | 515 (59.0%) |
| **MMSE** |  |  |  |
| Mean (sd) | 24.1 (4.3) | 26.2 (2.7) | 25.0 (3.8) |
| Range | 2 - 30 | 16 - 30 | 2 - 30 |
| Missing data | 2 | 2 | 4 |
| **APOE ε4 carrier/non carrier** |  |  |  |
| E2E2 | 0 (0.0%) | 2 (0.5%) | 2 (0.2%) |
| E2E3 | 11 (2.2%) | 53 (13.9%) | 64 (7.3%) |
| E2E4 | 10 (2.0%) | 7 (1.8%) | 17 (1.9%) |
| E3E3 | 141 (28.3%) | 239 (62.9%) | 378 (43.3%) |
| E3E4 | 236 (47.4%) | 65 (17.1%) | 298 (34.1%) |
| E4E4 | 92 (18.5%) | 6 (1.6%) | 98 (11.2%) |
| Missing | 8 (1.6%) | 8 (2.1%) | 16 (1.8%) |
| **Plasma p-tau217 Conc. (pg/mL)** |  |  |  |
| Median (sd) | 0.132 (0.09) | 0.046 (0.04) | 0.09 (0.08) |
| Range | 0.006 - 0.776 | 0.006 - 0.549 | 0.006 – 0.776 |
| **Amyloid Status Source** |  |  |  |
| CSF | 242 (48.6%) | 95 (25.0%) | 337 (38.6%) |
| PET | 253 (51.1%) | 283 (74.9%) | 536 (61.4%) |

#### p-Tau 217 Measurement in Plasma Samples

Figure 5 depicts p-Tau 217 sample results broken out by cohorts and subgroups. 100% of the samples were above the assay LoD and gave a reportable result. 99.5% of the samples were above the assay LLoQ and were thus quantifiable with acceptable precision. The median concentration of plasma p-Tau 217 was 2.87-fold higher in amyloid-positive study participants (amyloid negative 0.046 pg/mL, SD 0.04; amyloid positive 0.132 pg/mL, SD 0.09), p < 0.0001) and the differentiation between amyloid-positive and amyloid-negative study participants gave an overall AUC of 0.89 (0.87 - 0.92). There was a notable difference in discrimination between the ADC and Bio-Hermes cohorts, with the ADC training and validation subgroups yielding AUCs of 0.96 (0.94 – 0.99) and 0.93 (0.89 – 0.96) respectively vs. Bio-Hermes training and validation subgroups yielding AUCs of 0.89 (0.85 – 0.93) and 0.84 (0.78 – 0.89) respectively. This difference is considered further in the Discussion. A breakdown of performance metrics for various combinations of data sets is given in Table S4.

**Figure 5.**
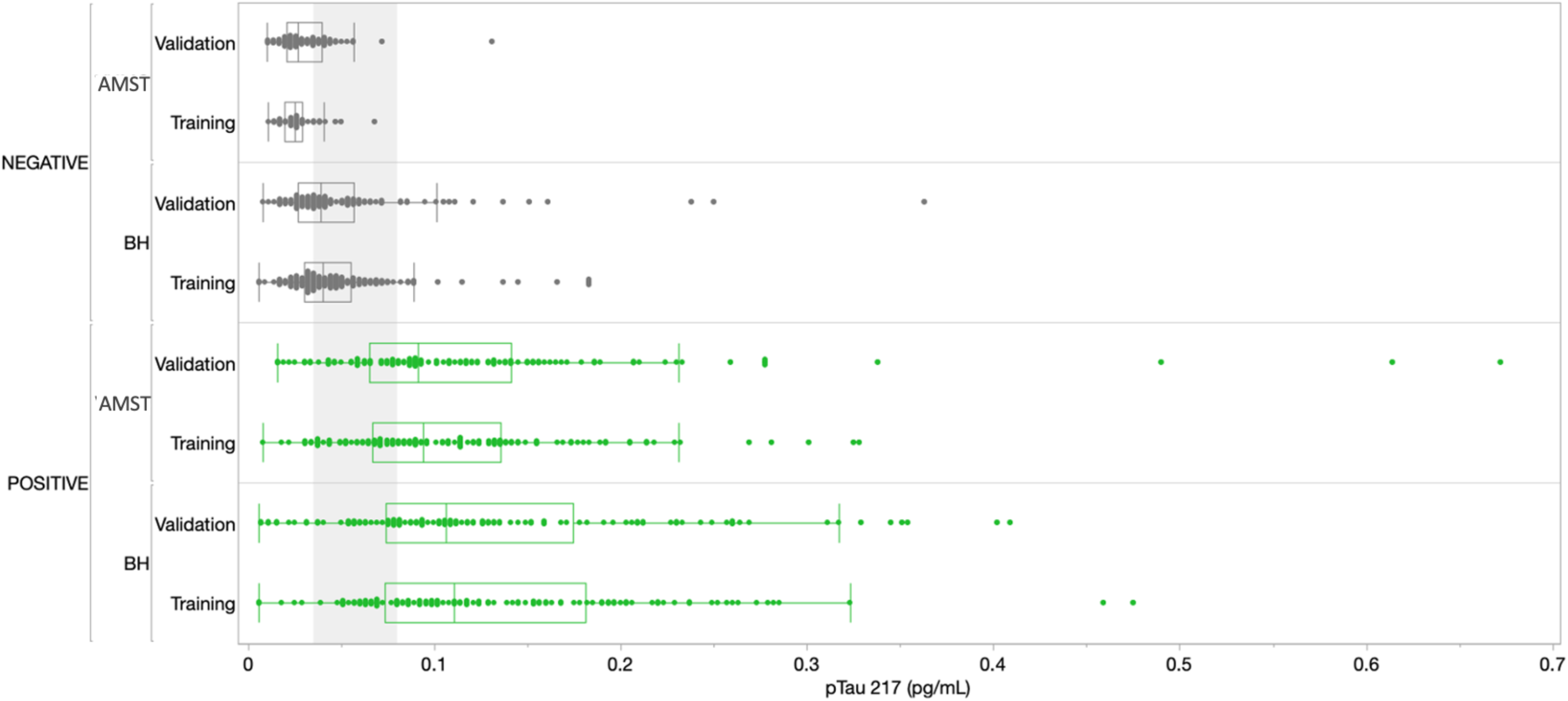
Distribution of results across all cohorts. Amyloid positives are depicted in green, amyloid negatives in grey. The grey shading corresponds to the ∼3O% intermediate zone of uncertainty between chosen lower and upper cutoffs of 0.04 and 0.09 pg/mL respectively (Figure 6).

In addition to the training and validation cohorts, subgroups of 50 each of diagnosed FTD and DLB cases were tested. Demographic and clinical details of these cases can be found in the Appendix Table A1. A proportion of these cases exhibited amyloid positivity (22% for FTD, 50% for DLB. Despite the limited statistical powering from the small sampling sizes, the data suggest amyloid detection accuracy statistically consistent with the validation cohort for detecting amyloid in DLB and FTD cases (85.0 and 87.5% respectively, Appendix Table A4). In addition, inclusion of all 100 cases to the validation cohort had no statistically significant effect on the performance of the test in classifying amyloid status (Appendix Figure A1).

#### Clinical Thresholds

In setting the lower and upper diagnostic thresholds, the objective was to maximize assay accuracy while minimizing the intermediate zone with an intended use population of objectively symptomatic individuals (MCI and AD). A simulation study was used to optimize the setting of the thresholds. Two threshold pairs representing the best balance were identified. Figure 6 depicts the clinical performance of the 0.035/0.080 pg/mL threshold pair, and a slightly higher 0.040/0.090 pg/mL threshold pair with respect to sensitivity, specificity, accuracy, and intermediate ranges across the subgroups. Note: sensitivity and specificity are reported here when excluding samples in the intermediate zone. In general, the lower candidate threshold pair favored sensitivity, while the higher threshold pair favored specificity (Figure 6A). Both candidate pairs gave similar performance for % intermediate zone and accuracy across the training subgroups. Generally, the wider spread of data observed in the Bio-Hermes cohort (Figure 5) contributed to a larger intermediate zone (∼36%) than with the ADC (∼25%). Combining cohorts gave an overall indeterminant range of ∼30% irrespective of the choice of threshold pairs. Overall, the higher threshold pair (0.04, 0.09 pg/mL) struck the best balance, yielding sensitivity, specificity, and accuracy >90% across the full data set, as well as PPV and NPV >90% with an amyloid prevalence of 50% representative of older patients with more concerning symptoms [7]. As reflected in Table 5, the validation subgroups had reduced estimated clinical performance relative to the training subgroups, in particular the Bio-Hermes validation subgroup. The main driver behind the difference was a higher number of false negatives among the Bio-Hermes validation subgroup (16) vs. the training subgroup (5). A deeper look revealed no obvious non-random demographic factors among the split, and the difference appeared to be a matter of chance. We summarized the performance of the p-Tau 217 assay across both training and validation cohorts, as reflected in Table 5. With the inclusion of all 873 patients across these two distinctly different independent cohorts, the test exhibited an overall accuracy of 90.7% excluding the intermediate range. The 30.9% intermediate range is mainly driven by the distribution spread introduced by the older, more diverse BioHermes cohort. PPV and NPV depend on the prevalence of amyloid positivity in the population being tested. Table S3 lists calculated PPV and NPV expected from populations with different disease prevalences, including the observed prevalence in this validation study (56%). In a population with low prevalence rates, such as among cognitively normal individuals or those with subjective cognitive decline (not yet validated), the Simoa p-Tau 217 test would exhibit a very high NPV (96-97%). Among patients with dementia where there is high prevalence of amyloid pathology, the test would exhibit a very high PPV (95%).

**Figure 6.**
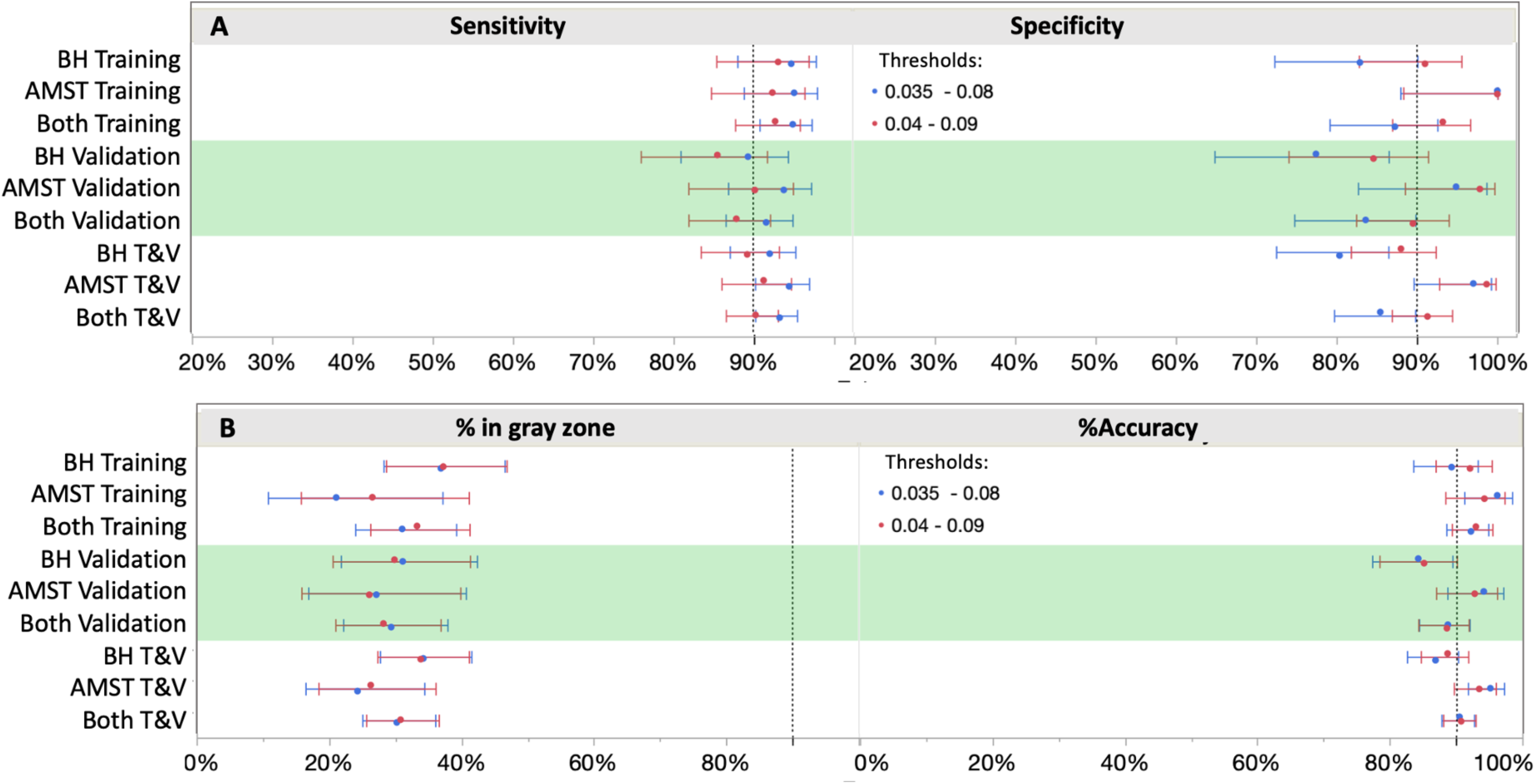
Clinical performance with two different cut-off scenarios. Shifting the cutoffs downward (blue) increased sensitivity, but at a higher cost to specificity. 0.04 and 0.09 pg/mL provided the best balance for both ruling out and ruling in with high confidence (≥90%), thus capturing both ends of the disease spectrum.

**Table 5:** Clinical performance with 0.04 and 0.09 pg/mL thresholds.

| Performance category | Amsterdam training<br>n=165 | Amsterdam validation<br>n=187 | BioHermes training<br>n=281 | BioHermes validation<br>n=240 | ADC + BioHermes<br>n=873 |
| --- | --- | --- | --- | --- | --- |
| AUC | 0.96 (0.94–0.99) | 0.93 (0.89–0.96) | 0.89 (0.85–0.93) | 0.84 (0.78–0.89) | 0.89 (0.87-0.92) |
| Sensitivity† | 92.4% (86.6-93.1%) | 90.2% (86.6-93.1%) | 93.1% (86.6-93.1%) | 85.6% (86.6-93.1%) | 90.3% (86.6-93.1%) |
| Specificity† | 100% (86.9-94.4%) | 97.8% (86.9-94.4%) | 91.0% (86.9-94.4%) | 84.6% (86.9-94.4%) | 91.3% (86.9-94.4%) |
| Accuracy†(NIA-AA) | 94.2% (88.0-92.9%) | 92.8% (88.0-92.9%) | 92.0% (88.0-92.9%) | 85.1% (88.0-92.9%) | 90.7% (88.0-92.9%) |
| Interm. range | 26.7% (25.7-36.7%) | 26.2% (25.7-36.7%) | 37.4% (25.7-36.7%) | 30.0% (25.7-36.7%) | 30.9% (25.7-36.7%) |

†Excluding samples in the intermediate range

#### Race and Ethnicity Analyses

While the ADC was primarily white/European, BioHermes represents an greater proportion of underserved racial/ethnicity (R/E) groups in the study cohort. The breakdown in R/E categories across all Bio-Hermes participants was approximately 74% white, 11% black/African American, 10% white/Latino, and 5% other/unknown. We attempted to discern whether there were any significant differences in test performance by examining p-Tau 217 levels for each R/E group in separate analyses. First, the proportions of clinical categories (MCI vs. AD) were not significantly different among the R/E groups (p = 0.0682). However, the amyloid positivity rate in the black/AA group was statistically lower as compared with other R/E groups (27.9% vs. ∼50%, Figure 7A). The likelihood-ratio test p-value was 0.0151 and an analysis of means of proportions showed that the black/AA group had a lower rate of positivity as compared to the overall rate of 46.8% across the study (Figure 7B). Importantly however, p-Tau 217 results did not differ significantly across R/E groups (Figure 8). Comparing all R/E pairs using a Tukey-Kramer multiple comparison indicated that the difference in p-Tau 217 results were among the largest between white and black/AA groups, but these differences did not reach statistical significance for either the amyloid positive subjects (mean difference 0.033 pg/mL, p = 0.3827) or the amyloid negative subjects (mean difference 0.013 pg/mL, p = 0.1137) (Table S6). Likewise, areas under the ROC curves ranged from 0.81 (0.67-0.96, (black/AA) to 0.89 (0.76-0.96, white Latino) (Figure S2).

**Figure 7.**
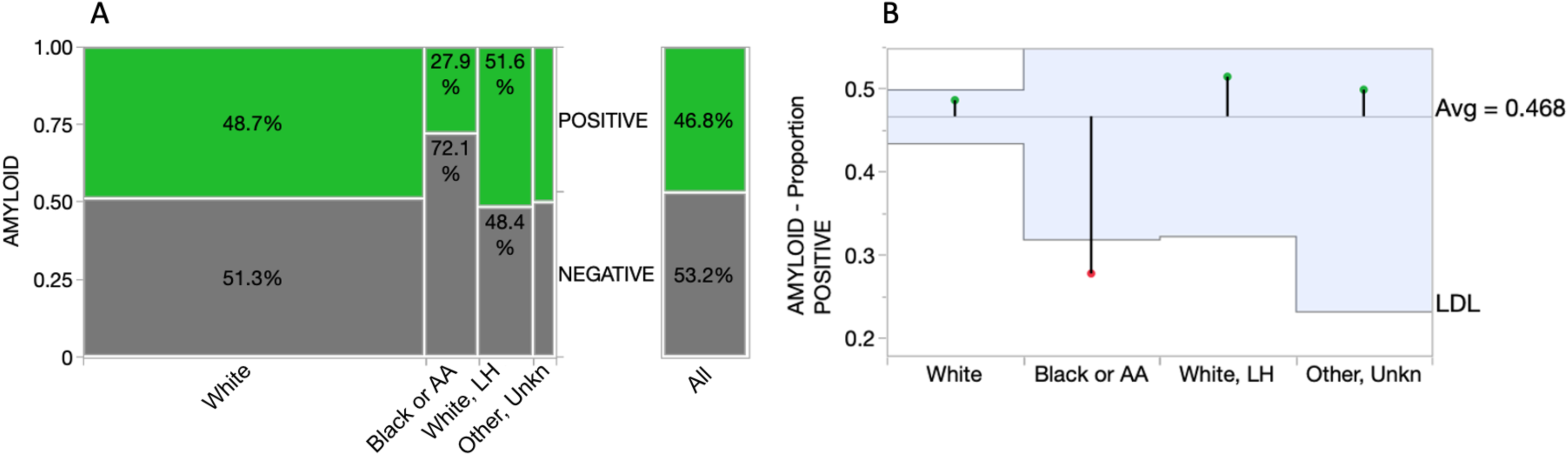
Comparison of amyloid positivity across racial/ethnic groups. Figure 7A: mosaic plot illustrating a signficiantly reduced percentage of amyloid positivity among Black/AA participants. Figure 7B: Analysis of Means for Proportions graph highlighting statistical significance of the lower proportion of amyloid positivity among Black/AA participants (p = 0.0151; box boundaries reflect 95% Cis). AA=African American, LH-Latino/Hispanic, LDL=lower decision limit.

**Figure 8.**
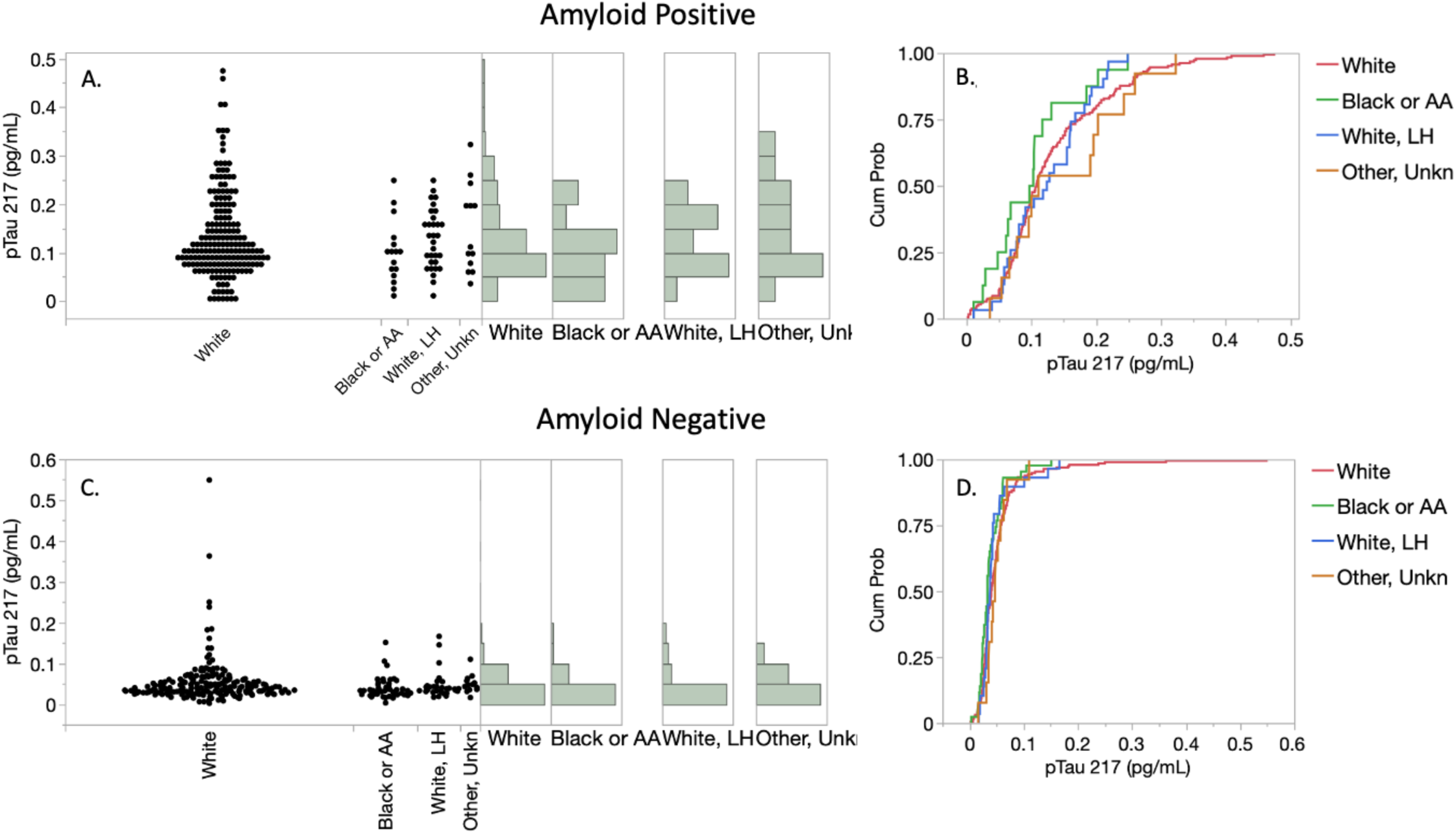
Distributions of p-Tau 217 results among racial/ethnic groups in the symptomatic Bio-Hermes cohort. Figures 8A, 8C: p-Tau 217 distributions. p-Tau 217 not differ significantly significantly across R/E groups, as reflected by cumulative distribution functions depicted in Figures 8B and 8D.

## Discussion

This report details the analytical and clinical validation of a simple, fully automated, and scalable digital immunoassay for accurate high sensitivity measurement of plasma p-Tau 217 that is suitable for routine clinical use. The test design and performance characteristics are aligned with the latest recommendations from expert groups on plasma test design and clinical performance capabilities needed to support confirmatory diagnostic use for identification of amyloid pathology in individuals with cognitive symptoms being evaluated for AD. In particular, use of two rather than one diagnostic threshold has been recommended for plasma p-Tau 217 [5, 7, 26], and the feasibility and diagnostic performance of 2-threshold plasma p-Tau 217 tests in clinical practice scenarios have been shown [26, 27]. Critically, these same expert sources unanimously reflect a consensus that a diagnostic accuracy of ≥90% (defined as the sum of correct results per comparator divided by all results [5]) is considered functionally equivalent to FDA-cleared CSF biomarker tests and suitable to enable a diagnostic use-case for a plasma AD biomarker. As shown in this report, the Simoa p-Tau 217 test achieves this high-performance standard across a well-powered clinical study diverse in participant demographics, geographies, comparator methods, clinical settings, and race/ethnicities. The high level of performance extends to clinical sensitivity and specificity (90.3, 91.3%, respectively) which is comparable to amyloid PET. For comparison, against gold standard postmortem neuropathology, qualitative amyloid PET has achieved reported sensitivities and specificities of 88-98% and 80-95% respectively [28, 29].

There was a notable difference in discrimination between the ADC and Bio-Hermes cohorts, with the ADC training and validation subgroups yielding higher AUC and clinical performance parameters vs. Bio-Hermes training and validation subgroups. There may be multiple reasons for this. One potential explanation is the greater racial/ethnic diversity of the Bio-Hermes cohort may have negatively impacted the diagnostic accuracy, although the racial/ethnic subgroup analysis didn’t reveal statistically significant differences (next section). Another potential explanation may be the presence of a larger number of comorbidities influencing the results in the older Bio-Hermes population. Detailed comorbidity information for the Bio-Hermes cohort was not available. It is also noted that Bio-Hermes utilized visual amyloid PET as the reference method, while the majority of the ADC utilized CSF biomarkers. It is unclear if visual amyloid PET may have introduced greater uncertainty in amyloid status than quantitative CSF classification. Yet another potential option for the observed differences may be the underlying methods by which the cohort individual subjects were assessed for clinical status. Also as reflected in Table 5, differences between training and validation subgroups despite randomization from the same sample cohorts suggests that with sampling sizes in the 240-280 range, fairly significant differences in clinical performance parameter estimates may emerge simply due to chance. This observation points to the importance of adequately powering validation studies to reliably determine the performance characteristics of an assay aimed at aiding Alzheimer’s diagnostics.

It is notable that the clinical validation reported here represents a culmination of several years of previously published data on the assay that was one of the first immunoassay-based tests for plasma p-Tau 217. The pedigree of the assay includes numerous cross sectional and longitudinal studies that were instrumental in helping establish the clinical validity of plasma p-Tau 217 for detection of amyloid and tau pathology. These studies include demonstration that the assay exhibits high accuracy as compared with amyloid and tau PET [3] and CSF biomarker status [30], detects p-Tau 217 elevation soon after brain Aβ levels begin to rise and prior to the rise in the earliest areas of tau accumulation [3], and outperforms p-Tau 181 and p-Tau 231 with stronger correlations and AUCs vs. amyloid and tau PET [31]. The assay has also been shown to exhibit accuracy indistinguishable from CSF p-Tau 217, whether early Alzheimer’s disease (A+T-) or more advanced Alzheimer’s disease (A+T+) [30], and predicts longitudinal cognitive changes as well as, or better than, amyloid or tau PET with a PPV > 90%, thus supporting its use as a substitute for PET in clinical trial enrollment [3, 32]. Among real-world clinical research studies, the assay achieved an AUC of 0.96 vs. FDA-cleared CSF Aβ42/p-Tau ratio test in discriminating amyloid positives from negatives in a clinically heterogenous population at a memory clinic [33]. While the clinical validation reported here was limited to individuals exhibiting cognitive symptoms, previous research studies show that the assay can accurately discriminate amyloid status among cognitively *unimpaired* older individuals [3, 34], indicating that the test may be clinically useful across the entire Alzheimer’s disease continuum with further validation.

A strength in the clinical validation study reported here was the use of two diverse independent cohorts representing different geographies (US/Europe), clinical settings (a tertiary clinic and a 17-site recruitment effort from under-represented communities), comparator methods (amyloid PET and CSF), diagnostic criteria (multi-disciplinary consensus and single primary provider assessment), prevalence’s (35.0% and 61.3% for MCI) and an emphasis on racial/ethnicity diversity (Bio-Hermes). As a result of this diversity in variables between the cohorts, differences in subgroup distributions were notable and the differences in diagnostic performance parameters likely reflect more real-world situations than might be the case with cohorts of more similar composition. The strong clinical performance characteristics achieved from the ADC, whether training or validation (accuracy 93-94%, specificity 98-100%, AUC 0.93-0.96) is consistent with previous reports of exceptional accuracy from a single clinical site [33] and could reflect a more homogeneous population (white/European) and highly vetted diagnostic criteria in a specialty setting. The relatively lower performance and lower amyloid positivity prevalence among MCI cases in the Bio-Hermes cohort may reflect the influence of racial/ethnic diversity, inconsistent diagnostic criteria across the sites, misdiagnosis prior to the availability of PET results, higher prevalence of comorbidities in an older population, and less accuracy in qualitative PET relative to CSF biomarkers. On this latter note, a comparison of qualitative PET classification with quantitative classification using a cutoff of 25 centiloids gave an 8% disagreement in classifications in the Bio-Hermes cohort putting a limitation on the maximum accuracy achievable with this cohort. These factors are also likely behind the breadth of the intermediate zone of 30.9%, whereas somewhere in the ∼20-25% range might have been anticipated as reported for other immunoassays. Supporting this, a recent comparison between the Simoa p-Tau 217 described in this report and an alternative Simoa p-Tau 217 assay with different antibodies showed the two assays gave statistically indistinguishable clinical performance [35], yet a second report on the alternative assay suggested a 20% intermediate zone was suitable for the cohorts tested [36]. The cohorts tested here may be more representative of real-world variables and yield a diagnostic performance more realistic for an immunoassay used in a natural population.

p-Tau 217 is a low abundance protein in plasma, making it challenging to reliably measure in non-diseased and early/prodromal diseased individuals. As AD pathophysiology progresses, levels of plasma p-Tau 217 elevate, easing the analytical challenge of its measurement. As reported here, the digital Simoa p-Tau217 immunoassay with single femtogram/mL sensitivity makes it possible to reliably measure p-Tau 217 in all subjects, whether diseased or healthy. In contrast, a well published electrochemiluminescence immunoassay method was unable to detect plasma p-Tau 217 in 13% of samples tested [37], and a mass spectrometry-based algorithmic method combining p-Tau 217 with non-phosphorylated tau for a %p-Tau ratio was unable to detect p-Tau 217 in 27% of the samples tested [38]. In the latter case, artificial p-Tau 217 values of ½ the LoD (1.3 pg/mL) were imputed into the algorithm to obtain a result. These scenarios make it more difficult to interpret apparent fold changes in median values between AD and control populations (often used to compare performance between assay methods) and to perform correlation analyses between methods. Additionally, very low abundance p-Tau 217 is expected to be common in individuals with low amyloid burden, limiting the potential for tracking the biomarker longitudinally, for example in asymptomatic individuals such as those with SCD. Finally, methods with inadequate analytical sensitivity may not be suitable for precise assessment of biomarker status at or near a lower diagnostic threshold as would be needed for high confidence in ruling out the presence of amyloid pathology. Such tests may be limited to identifying only patients with high amyloid burden and sufficiently high plasma p-Tau 217 to support use as a rule-in test, potentially decreasing the benefit of a plasma biomarker test to reduce the number of lumbar punctures or PET scans during diagnostic evaluation, as well as the potential benefit of reducing the flow of referrals from primary care to specialty care.

A goal of the Bio-Hermes study was the participation of at least 20% of the cohort from traditionally underserved population groups. As was reported previously, the proportion of participants who were positive for amyloid by PET was statistically lower for non-Hispanic Blacks compared to non-Hispanic Whites [20]. This finding held for the symptomatic clinical groups studied here. Likewise, the proportions of amyloid positive subjects did not differ among Hispanics and non-Hispanic Whites. It remains unclear why there was a lower prevalence of amyloid positivity among non-Hispanic blacks in these studies, but perhaps it is related to differences in education levels and cognitive scoring that were found to be significant [20] combined with a tendency to over-diagnose in the absence of PET results (which were obtained only after diagnosis). Also consistent was our finding among symptomatic individuals that differences in plasma p-Tau 217 between R/E groups did not attain statistical significance. As previously reported, p-Tau 181 and amyloid-beta ratio also did not differ between R/E groups [20]. Importantly, the ideal plasma p-Tau 217 cutoffs for identifying amyloid status did not significantly differ between R/E groups in this study. It seems likely that discerning R/E differences requires larger and/or more diverse cohorts with greater power than the Bio-Hermes cohort provides. Nonetheless, it is reassuring that R/E differences in plasma p-Tau 217 seem to be absent to fairly minor. Differences in diagnostic performance based on sex, age, and apoE4 carriership were also found not to be significant (not shown). However, even small differences could have a significant impact when used for large-scale screening of populations. As the impact of demographic variables is explored more fully, guidance could be developed regarding the interpretation of p-Tau 217 test results in the context of these variables. Taken together the data here suggest that the results of the Simoa p-Tau 217 test can be similarly interpreted across different ethnicities, ages, sexes, and apoE4 genotypes.

The study is not without limitations. While the ADC represents tertiary care clinical practice and reflects all comers to the clinic without exclusions, the R/E composition was more limited to primarily to individuals of white European descent. On the other hand, the Bio-Hermes cohort was aimed at greater diversity, but the participants were recruited and evaluated at clinical research entities in a similar manner to therapeutic trial enrollment rather than at primary or secondary clinics. The Bio-Hermes study enrollment included various exclusions, including prior history of cancer, psychiatric conditions, recent alcohol dependence, other non-AD factors that could contribute to cognitive symptoms (e.g., bladder infection), underweight, potential competing neurological disorders, etc. In the BioHermes cohort, comorbidities such as renal function, cardiovascular disease, and brain trauma were not captured or controlled for. While the potential for co-morbidities to affect plasma biomarker concentrations has been a topic of considerable discussion in the context of clinical implementation of blood tests for AD, recent data suggest that the effect of what is generally considered the most impactful comorbidity--chronic kidney disease-

-may not be clinically meaningful for correct classification of amyloid status using plasma p-Tau 217 [39]. Nonetheless, the potential for a higher prevalence of undocumented co-morbidities in the Bio-Hermes cohort composed of older participants and the emphasis on underserved minority participants may have been a contributing factor to the weaker diagnostic performance with this cohort relative to the ADC. Additional studies are ongoing to examine the effect of comorbidities on the Simoa p-Tau 217 test.

## Conclusions

The Simoa p-Tau 217 blood test was clinically validated across two diverse independent cohorts of individuals with cognitive impairment. The test employs a two-cutoff design according to the high performance criteria that has been recently recommended for diagnostic confirmatory use case, with an overall accuracy vs. amyloid PET and CSF of >90%, and sensitivity and specificity >90%. This two-cutoff design led to an intermediate zone of ∼30% with the cohorts studied here. At an amyloid prevalence of 50% reflecting mild cognitive impairment the test also exhibited >90% PPV and NPV. The test was analytically validated and shown to deliver single femtogram/mL sensitivity, enabling the measurement of plasma p-Tau 217 in all individuals tested. These results demonstrate that this Simoa plasma p-Tau 217 test as validated under CLIA is suitable for clinical use.

## Supporting information

Appendix- FTD & DLB analyses

## Data Availability

Data produced in the present study are available upon reasonable request to the authors

## Acknowledgements

Inge M.W. Verberk is supported by grants of the Alzheimer’s Association, Health∼Holland and Amsterdam UMC. The chair of Wiesje M. van der Flier is supported by the Pasman stichting. Charlotte E. Teunissen are recipients of TAP-dementia (www.tap-dementia.nl), receiving funding from ZonMw (#10510032120003) in the context of Onderzoeks programma Dementie, part of the Dutch National Dementia Strategy. Wiesje M. van der Flier and Charlotte E. Teunissen are recipients of ABOARD, which is a public-private partnership receiving funding from ZonMW (#73305095007) and Health∼Holland, Topsector Life Sciences & Health (PPP-allowance; #LSHM20106). Alzheimer Nederland. Alzheimer Center Amsterdam is supported by Stichting Alzheimer Nederland and Stichting Steun Alzheimercentrum Amsterdam. Charlotte E. Teunissen further received grants of the European Commission (Marie Curie International Training Network, grant agreement No 860197 (MIRIADE), Innovative Medicines Initiatives 3TR (Horizon 2020, grant no 831434) EPND (IMI 2 Joint Undertaking (JU), grant No. 101034344), and JPND (bPRIDE), National MS Society (Progressive MS Alliance), Alzheimer Drug Discovery Foundation, Alzheimer Association, Health Holland, the Dutch Research Council (ZonMW), the Selfridges Group Foundation, and Alzheimer Netherlands. The Amsterdam Dementia Cohort clinical database structure was developed with funding from Stichting Dioraphte.

## Ethics Statement

All studies were properly consented and approved by applicable ethics committees. The studies were conducted in accordance with local legislation and institutional requirements.

