## Supplementary Figures & Tables for "Analytical and Clinical Validation of a High Accuracy Fully Automated Digital Immunoassay for Plasma Phospho-Tau 217 for Clinical Use in Detecting Amyloid Pathology"

**Figure S1: Age Distributions of Bio-Hermes and Amsterdam Cohort**

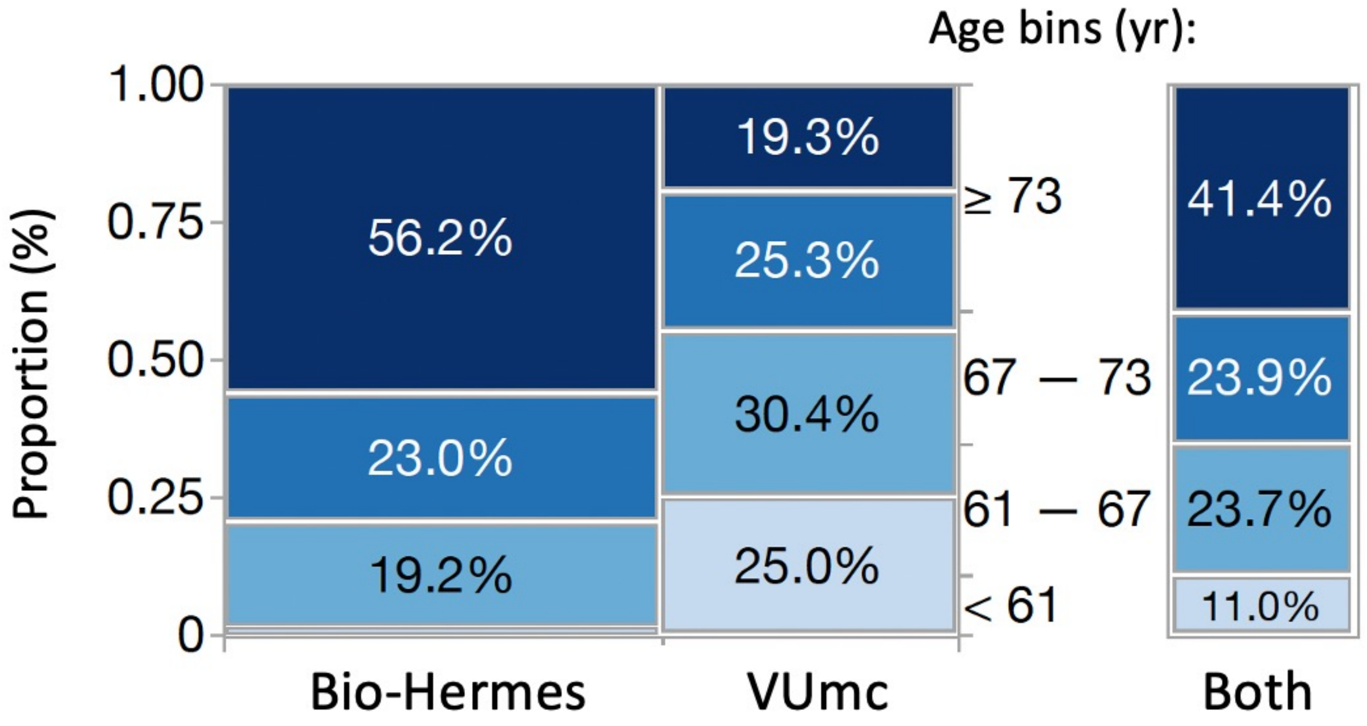


**Figure S1**: Proportion of participant ages by bins across the two cohorts.

### Figure S2: ROC Curves Across Racial/Ethnic Groups

Bio-Hermes symptomatic groups cohort only

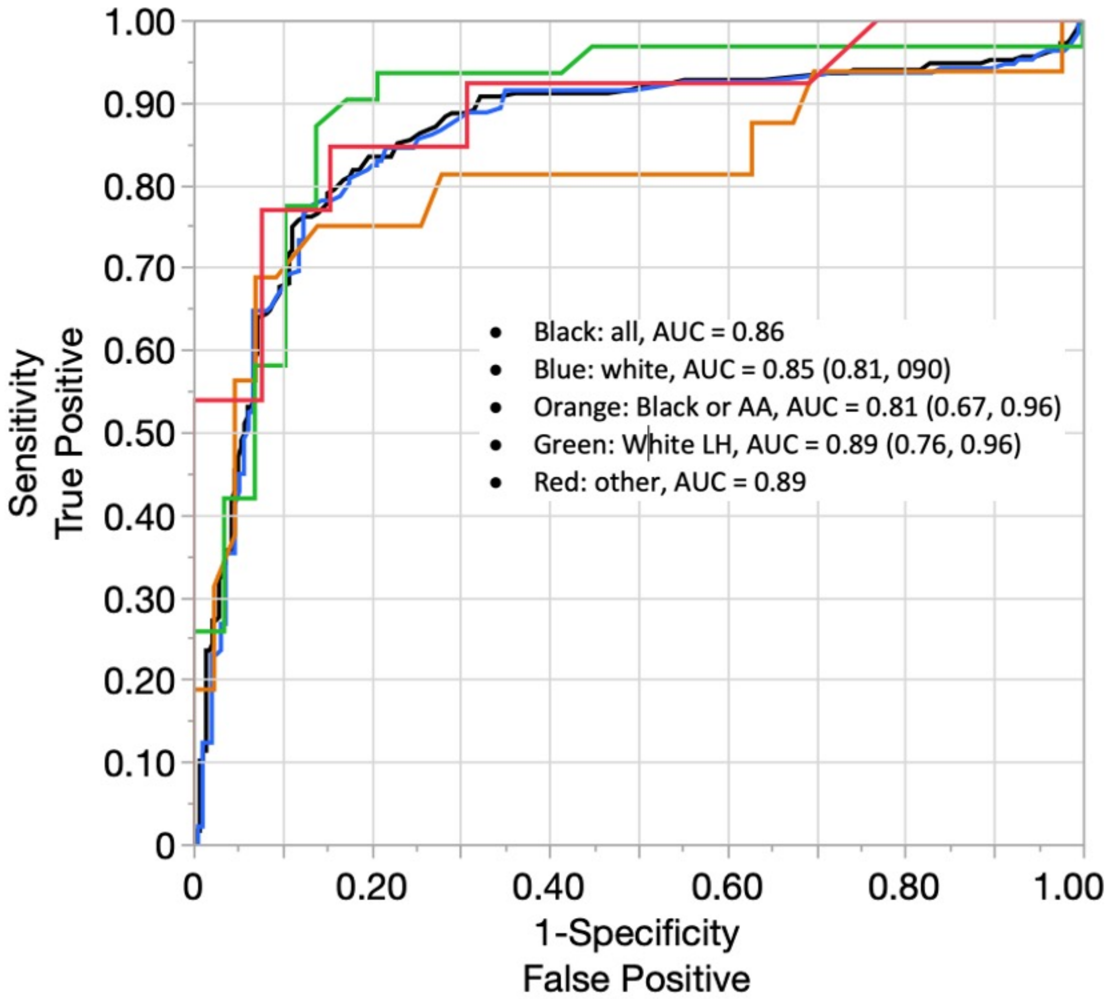


**Figure S2**: ROC curves as predictors of amyloid status (with 95% CI's)

### Table S1:
### Demographic characteristics by cohort

| | BH N = 521 | VUMC N = 352 | All N =873 |
|---|---|---|---|
| **Age** | | | |
| Mean (sd) | 73.2 (6.6) | 65.4 (7.7) | 70.1 (8.0) |
| Range | 59 – 85 | 43 – 83 | 43 - 85 |
| **Sex** | | | |
| Female | 277 (53.2%) | 162 (46.0%) | 439 (50.3%) |
| **Race** | | | |
| American Indian or Alaska Native | 1 (0.2%) | 0 (0.0%) | 1 (0.1%) |
| Asian | 10 (1.9%) | 0 (0.0%) | 10 (1.5%) |
| Black or African American | 60 (11.5%) | 0 (0.0%) | 60 (8.9%) |
| Native Hawaiian or other Pacific Islander | 1 (0.2%) | 0 (0.0%) | 1 (0.1%) |
| Other/Unknown | 5 (1.0%) | 14 (9.0%) | 19 (2.8%) |
| White | 444 (85.2%) | 142 (91.0%) | 586 (86.6%) |
| Missing† <br>†Not included in %s | 0 | 196 | 196 |
| **Ethnicity** | | | |
| Hispanic or Latino | 68 (13.1%) | 0 (0.0%) | 68 (7.8% ) |
| Not Hispanic or Latino | 446 (85.6%) | 0 (0.0%) | 446 (51.1%) |
| Not Reported | 7 (1.34%) | 0 (0.0%) | 7 (0.8% ) |
| Unknown | 0 (0.0%) | 352 | 352 (40.3%) |
| **Diagnosis** | | | |
| AD | 0 (0.0%) | 123 (34.9%) | 123 (14.1%) |
| Probable AD | 235 (45.1%) | 0 (0.0%) | 235 (26.9%) |
| MCI | 286 (54.9%) | 229 (65.1%) | 515 (59.0%) |
| **MMSE** | | | |
| Mean (sd) | 24.1 (4.3) | 26.2 (2.7) | 25.0 (3.8) |
| Range | 2 - 30 | 16 - 30 | 2 - 30 |
| Missing data | 2 | 2 | 4 |
| **APOE e4 carrier/non carrier** | | | |
| E2E2 | 1 (0.2%) | 1 (0.3%) | 2 (0.2%) |
| E2E3 | 49 (9.4%) | 15 (4.3%) | 64 (7.3%) |
| E2E4 | 13 (2.5%) | 4 (1.1%) | 17 (1.9%) |
| E3E3 | 259 (49.7%) | 119 (33.8%) | 378 (43.3%) |
| E3E4 | 166 (31.9%) | 132 (37.5%) | 298 (34.1%) |
| E4E4 | 33 (6.3%) | 65 (18.5%) | 98 (11.2%) |
| Missing | 0 (0.0%) | 16 (4.5%) | 16 (1.8%) |
| **Plasma p-tau217 Conc. (pg/mL)** | | | |
| Mean (sd) | 0.09 (0.08) | 0.10 (0.09) | 0.09 (0.08) |
| **Amyloid Status** | | | |
| CSF | 0 (0.0%) | 337 (95.7%) | 337 (38.6%) |
| PET | 521 (100.0%) | 15 (4.3%) | 536 (61.4%) |

**Table S1**: Amyloid classification based on amyloid PET or CSF biomarkers. Abbreviations: MCI = mild cognitive impairment; MMSE = Mini-Mental State Examination; PET = positron emission tomography.

#### Table S2: Amyloid Prevalence by Diagnostic Category

| | | AMYLOID | | | | | |
|---|---|---|---|---|---|---|---|
| | | POSITIVE | | NEGATIVE | | All | |
| Source | Group Simple | N | Row % | N | Row % | N | Row % |
| BH | Probable AD | 144 | 61.28% | 91 | 38.72% | 235 | 100.00% |
| | MCI | 100 | 34.97% | 186 | 65.03% | 286 | 100.00% |
| | All | 244 | 46.83% | 277 | 53.17% | 521 | 100.00% |
| VUMC | AD | 122 | 99.19% | 1 | 0.81% | 123 | 100.00% |
| | MCI | 129 | 56.33% | 100 | 43.67% | 229 | 100.00% |
| | All | 251 | 71.31% | 101 | 28.69% | 352 | 100.00% |
| All | All | 495 | 56.70% | 378 | 43.30% | 873 | 100.00% |

#### Table S3: PPV and NPV of Simoa p-Tau 217 assay by prevalence

| Population | Prevalence | PPV | NPV |
|---|---|---|---|
| Cognitively normal older adults | 20.0% | 72.2% | 97.4% |
| Subjective cognitive decline | 30.0% | 81.6% | 95.6% |
| Mild cognitive impairment | 50.0% | 91.2% | 90.4% |
| Prevalence in this study | 56.7% | 93.1% | 87.8% |
| Dementia population | 65.0% | 95.1% | 83.5% |

Typical prevalences for different populations, and the prevalence observed in the study reported here. Abbreviations: PPV, positive predictive value; NPV, negative predictive value.

#### Table S4: **Performance Metrics with 95% CI for Various Combinations of Data Sets**

| Data Set | Measures | Est | LCL | UCL |
|---|---|---|---|---|
| BH Training | Prevalence | 43.8% | 35.3% | 52.6% |
| | False Negative Rate | 4.9% | 0.3% | 44.7% |
| | False Positive Rate | 5.1% | 0.4% | 38.8% |
| | % in Indeterminant Range | 37.4% | 28.7% | 46.9% |
| | Accuracy | 92.0% | 86.8% | 95.3% |
| | Sensitivity | 93.1% | 85.4% | 96.9% |
| | Specificity | 91.0% | 82.8% | 95.5% |
| AMST Training | Prevalence | 78.8% | 71.0% | 84.9% |
| | False Negative Rate | 5.4% | 0.4% | 41.9% |
| | False Positive Rate | 0.0% | . | . |
| | % in Indeterminant Range | 26.7% | 15.9% | 41.2% |
| | Accuracy - Ind | 94.2% | 88.3% | 97.2% |
| | Sensitivity | 92.4% | 84.7% | 96.4% |
| | Specificity | 100.0% | 88.3% | 100.0% |
| Both Training | Prevalence | 56.7% | 50.6% | 62.7% |
| | False Negative Rate | 5.1% | 0.7% | 30.1% |
| | False Positive Rate | 4.1% | 0.3% | 37.7% |
| | % in Indeterminant Range | 33.4% | 26.3% | 41.3% |
| | Accuracy - Ind | 92.9% | 89.3% | 95.4% |
| | Sensitivity | 92.7% | 87.7% | 95.8% |
| | Specificity | 93.2% | 86.9% | 96.6% |
| BH Validation | Prevalence | 50.4% | 41.6% | 59.2% |
| | False Negative Rate | 10.7% | 2.4% | 37.0% |
| | False Positive Rate | 10.1% | 2.1% | 37.5% |
| | % in Indeterminant Range | 30.0% | 20.6% | 41.4% |
| | Accuracy - Ind | 85.1% | 78.4% | 90.0% |
| | Sensitivity | 85.6% | 76.0% | 91.7% |
| | Specificity | 84.6% | 74.0% | 91.4% |
| AMST Validation | Prevalence | 64.7% | 55.9% | 72.6% |
| | False Negative Rate | 7.4% | 1.0% | 39.4% |
| | False Positive Rate | 1.5% | 0.0% | 80.0% |
| | % in Indeterminant Range | 26.2% | 16.0% | 39.9% |
| | Accuracy - Ind | 92.8% | 86.9% | 96.1% |
| | Sensitivity | 90.2% | 81.9% | 94.9% |
| | Specificity | 97.8% | 88.5% | 99.6% |
| Both Validation | Prevalence | 56.7% | 50.4% | 62.8% |
| | False Negative Rate | 9.1% | 2.5% | 27.8% |
| | False Positive Rate | 7.0% | 1.2% | 32.5% |
| | % in Indeterminant Range | 28.3% | 21.1% | 36.9% |
| | Accuracy - Ind | 88.6% | 84.2% | 91.8% |
| | Sensitivity | 87.9% | 82.0% | 92.1% |
| | Specificity | 89.5% | 82.4% | 94.0% |
| BH Training & Validation | Prevalence | 46.8% | 40.7% | 53.1% |
| | False Negative Rate | 7.8% | 1.8% | 28.0% |
| | False Positive Rate | 7.2% | 1.6% | 26.6% |
| | % in Indeterminant Range | 34.0% | 27.4% | 41.2% |
| | Accuracy - Ind | 88.7% | 84.6% | 91.8% |
| | Sensitivity | 89.3% | 83.5% | 93.2% |
| | Specificity | 88.0% | 81.8% | 92.3% |
| AMST Training & Validation | Prevalence | 71.3% | 65.4% | 76.5% |
| | False Negative Rate | 6.4% | 1.2% | 28.5% |
| | False Positive Rate | 1.0% | 0.0% | 79.8% |
| | % in Indeterminant Range | 26.4% | 18.5% | 36.2% |
| | Accuracy - Ind | 93.4% | 89.6% | 95.9% |
| | Sensitivity | 91.3% | 86.1% | 94.7% |
| | Specificity | 98.7% | 92.8% | 99.8% |
| **Both Training & Validation** | **Prevalence** | **56.7%** | **52.3%** | **61.0%** |
| | **False Negative Rate** | **7.1%** | **2.2%** | **20.4%** |
| | **False Positive Rate** | **5.6%** | **1.1%** | **23.8%** |
| | **% in Indeterminant Range** | **30.9%** | **25.7%** | **36.7%** |
| | **Accuracy** | **90.7%** | **88.0%** | **92.9%** |
| | **Sensitivity** | **90.3%** | **86.6%** | **93.1%** |
| | **Specificity** | **91.3%** | **86.9%** | **94.4%** |

#### Table S5: Tukey-Kramer Comparisons of Pairwise Differences in Mean p-Tau 217 Values Across Racial/Ethnic Groups

##### Amyloid Positives

| Level | - Level | Difference | Std Err Dif | Lower CL | Upper CL | p-Value |
|---|---|---|---|---|---|---|
| White | Black or AA | 0.0129517 | 0.0081627 | -0.003118 | 0.0290210 | 0.1137 |
| Other, Unkn | Black or AA | 0.0101020 | 0.0153279 | -0.020073 | 0.0402769 | 0.5104 |
| White, LH | Black or AA | 0.0071524 | 0.0116367 | -0.015756 | 0.0300606 | 0.5393 |
| White | White, LH | 0.0057993 | 0.0096416 | -0.013181 | 0.0247800 | 0.5480 |
| Other, Unkn | White, LH | 0.0029496 | 0.0161640 | -0.028871 | 0.0347705 | 0.8553 |
| White | Other, Unkn | 0.0028497 | 0.0138742 | -0.024463 | 0.0301629 | 0.8374 |

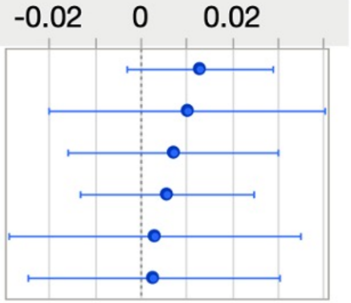


##### Amyloid Negatives

| Level | - Level | Difference | Std Err Dif | Lower CL | Upper CL | p-Value |
|---|---|---|---|---|---|---|
| White | Black or AA | 0.0129517 | 0.0081627 | -0.003118 | 0.0290210 | 0.1137 |
| Other, Unkn | Black or AA | 0.0101020 | 0.0153279 | -0.020073 | 0.0402769 | 0.5104 |
| White, LH | Black or AA | 0.0071524 | 0.0116367 | -0.015756 | 0.0300606 | 0.5393 |
| White | White, LH | 0.0057993 | 0.0096416 | -0.013181 | 0.0247800 | 0.5480 |
| Other, Unkn | White, LH | 0.0029496 | 0.0161640 | -0.028871 | 0.0347705 | 0.8553 |
| White | Other, Unkn | 0.0028497 | 0.0138742 | -0.024463 | 0.0301629 | 0.8374 |

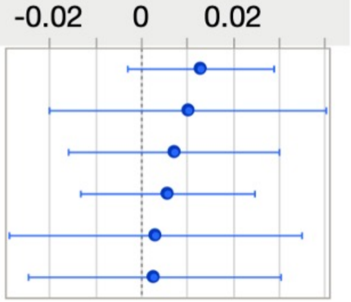
