## Appendix- FTD & DLB analyses for "Analytical and Clinical Validation of a High Accuracy Fully Automated Digital Immunoassay for Plasma Phospho-Tau 217 for Clinical Use in Detecting Amyloid Pathology"

Supplementary analyses: Accuracy of Simoa p-Tau 217 (LucentAD p-Tau 217) for dementia with Lewy bodies (DLB) and frontal temporal dementia (FTD) cases.

50 each of cases diagnosed with DLB and FTD were tested in the assay with optimized diagnostic thresholds. A proportion of these samples were also amyloid positive, and the accuracy of the test for detection of amyloid in these mixed pathology cases was characterized. Demographic and clinical characteristics of these samples are summarized in Table A1.

**Table A1: Demographic and clinical characteristics of DLB and FTD samples from the ADC**

| | DLB | FTD |
|---|---|---|
| n | 50 | 50 |
| Age (mean, SD) | 68.14 | 62.42 |
| Sex (male, %) | 41 (82.0) | 28 (56.0) |
| APOE carrier = yes (%) | 26 (53.1) | 12 (26.1) |
| MMSE (mean, SD) | 22.22 (4.84) | 24.29 (4.60) |
| CSF Abeta42 (mean, SD) | 772.22 (240.93) | 943.77 (280.47) |
| CSF p-Tau (mean, SD) | 51.72 (21.18) | 48.35 (21.89) |
| CSF Tau (mean, SD) | 377.46 (203.08) | 399.82 (234.98) |
| Amyloid positive by CSF (%) | 50% | 22% |

p-Tau 217 results compared with CSF amyloid status for the DLB and FTD cases are summarized in Tables A2 and A3 below.

**Table A2: 2 x 3 Table for DLB cases**

| | | **p-Tau 217 Result** (Intermediate zone: 0.04 - 0.09 pg/mL) | | | | | | | |
|---|---|---|---|---|---|---|---|---|---|
| | | **Low Risk** | | **Intermediate** | | **High Risk** | | **All** | |
| **Clinical population** | **Amyloid Status** | **N** | **Row %** | **N** | **Row %** | **N** | **Row %** | **N** | **Row %** |
| DLB | Positive | 0 | 0.0% | 20 | 80.0% | 5 | 20.0% | 25 | 100.0% |
| | Negative | 12 | 48.0% | 10 | 40.0% | 3 | 12.0% | 25 | 100.0% |
| | All | 12 | 24.0% | 30 | 60.0% | 8 | 16.0% | 50 | 100.0% |

**Table A3: 2 x 3 Table for FTD cases**

| | | **p-Tau 217 Result** (Intermediate zone: 0.04 - 0.09 pg/mL) | | | | | | | |
|---|---|---|---|---|---|---|---|---|---|
| | | **Low Risk** | | **Intermediate** | | **High Risk** | | **All** | |
| **Clinical population** | **Amyloid Status** | **N** | **Row %** | **N** | **Row %** | **N** | **Row %** | **N** | **Row %** |
| FTD | Positive | 2 | 18.2% | 4 | 36.4% | 5 | 45.5% | 11 | 100.0% |
| | Negative | 23 | 59.0% | 14 | 35.9% | 2 | 5.12% | 39 | 100.0% |
| | All | 25 | 50.0% | 18 | 36.0% | 7 | 14.0% | 50 | 100.0% |

Clinical performance metrics obtained for the DLB and FTD cases are summarized in Table A4:

**Table A4: Performance metrics with 95% CI for DLB and FTD cases**

| Data Set | Measures | Est | LCL | UCL |
|---|---|---|---|---|
| DLB | Amyloid prevalence | 50.0% | 36.6% | 63.4% |
| | False Negative Rate | 0.0% | 0.0% | 13.3% |
| | False Positive Rate | 12.0% | 4.2% | 30.0% |
| | % in intermediate zone | 60.0% | 46.2% | 72.4% |
| | Accuracy (- Int) | 85.0% | 64.0% | 94.8% |
| | Sensitivity (- Int) | 100.0% | 56.6% | 100.0% |
| | Specificity (- Int) | 80.0% | 54.8% | 93.0% |
| FTD | Amyloid prevalence | 22.0% | 12.8% | 35.2% |
| | False Negative Rate | 18.2% | 5.1% | 47.7% |
| | False Positive Rate | 5.1% | 1.4% | 16.9% |
| | % in intermediate zone | 36.0% | 24.1% | 49.9% |
| | Accuracy (- Int) | 87.5% | 71.9% | 95.0% |
| | Sensitivity (- Int) | 71.4% | 35.9% | 91.8% |
| | Specificity (- Int) | 92.0% | 75.0% | 97.8% |

Despite the limited statistical powering from the small sampling sizes, the data suggest amyloid detection accuracy statistically consistent with the validation cohort for detecting amyloid in DLB and FTD cases. It is noted that although 100% sensitivity was observed in DLB cases, a majority (30) fell into the intermediate zone, suggesting a relatively weak amyloid signal.

The impact of inclusion of these non-AD and co-pathology cases into the validation cohort was assessed. Two different incidence levels were examined: “typical” percentages as reported in memory clinics, and “high” levels as might be encountered with under-diagnoses and among a younger population with high percentages of FTD. Table A5 summarizes the percentages that were included:

**Table A5: Percentage of DLB and FTD cases included in validation cohort**

| Dx category | Typical | High |
|---|---|---|
| DLB | 4.6%[A1] | 9.5% |
| FTD | 7.7%[A2-5] | 9.5%[A-6] |

The effect of the addition of up to 100 non-AD and AD co-pathology cases is depicted in Figure A1 below.

**Figure A1: Effect of the addition of DLB and FTD samples on performance metrics**

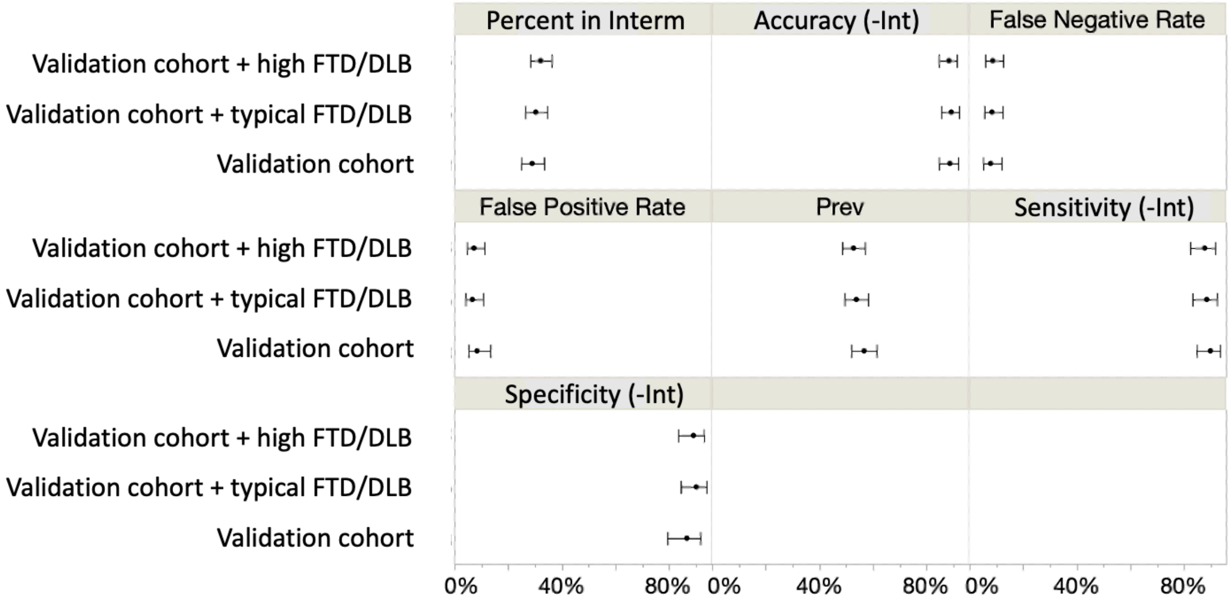


While the 30 DLB cases in the intermediate zone increased the overall validation cohort intermediate zone from 28% to 32%, there was no significant difference in the performance of the test in classifying amyloid status with up to 19% of non-AD and co-pathology cases added to the cohort.
